# Meta-Analysis Reveals the Vaginal Microbiome is a Better Predictor of Earlier Than Later Preterm Birth

**DOI:** 10.1101/2022.09.26.22280389

**Authors:** Caizhi Huang, Craig Gin, Jennifer Fettweis, Betsy Foxman, Bizu Gelaye, David A. MacIntyre, Akila Subramaniam, William Fraser, Negar Tabatabaei, Benjamin Callahan

## Abstract

High-throughput sequencing measurements of the vaginal microbiome have yielded intriguing potential relationships between the vaginal microbiome and preterm birth (PTB; live birth prior to 37 weeks of gestation). However, results across studies have been inconsistent. Here we perform an integrated analysis of previously published datasets from 12 cohorts of pregnant women whose vaginal microbiomes were measured by 16S rRNA gene sequencing. Of 1926 women included in our analysis, 568 went on to deliver prematurely. Substantial variation between these datasets existed in their definition of preterm birth, characteristics of the study populations, and sequencing methodology. Nevertheless, a small group of taxa comprised a vast majority of the measured microbiome in all cohorts. We trained machine learning (ML) models to predict PTB from the composition of the vaginal microbiome, finding low to modest predictive accuracy (0.28-0.79). Predictive accuracy was typically lower when ML models trained in one dataset predicted PTB in another dataset. Earlier preterm birth (<32 weeks, <34 weeks) was more predictable from the vaginal microbiome than late preterm birth (34 - 37 weeks), both within and across datasets. Integrated differential abundance analysis revealed a highly significant negative association between *L. crispatus* and PTB that was consistent across almost all studies. The presence of the majority (18 out of 25) of genera was associated with a higher risk of PTB, with *L. iners, Prevotella*, and *Gardnerella* showing particularly consistent and significant associations. Some example discrepancies between studies could be attributed to specific methodological differences, but not most study-to-study variations in the relationship between the vaginal microbiome and preterm birth. We believe future studies of the vaginal microbiome and PTB will benefit from a focus on earlier preterm births, and improved reporting of specific patient metadata shown to influence the vaginal microbiome and/or birth outcomes.

## Introduction

Preterm birth (PTB), defined as live birth prior to 37 complete weeks of gestation, is the primary cause of neonatal morbidity and mortality worldwide with an average PTB prevalence of around 11% (Chawanpaiboon et al.,2019; Manuck et al.,2016). However, our current understanding of PTB is limited with no clear causative factor for the majority of PTBs (Ferrero et al.,2016). One of the known risk factors of PTB is bacterial-related inflammation in gestational tissue (Donders et al.,2009;Goldenberg et al.,2008), including bacterial vaginosis (BV) – a polymicrobial alteration of the vaginal microbiome characterized by depletion of *Lactobacillus* species and overgrowth of typically strict anaerobes (Onderdonk et al.,2016).

In the past decade, the development of high-throughput sequencing technologies has reformed the study of the human microbiome. High throughput sequencing of PCR-amplified marker genes (e.g., 16S rRNA gene), and shotgun metagenomic sequencing of total sample DNA are now widely used to measure the composition and functional potential of whole microbial communities. To date, at least 15 studies have used high-throughput sequencing to investigate the link between the vaginal microbiome and PTB, or preterm premature rupture of membranes (PPROM), which precedes between 30-40% of all PTB cases. All these studies employed a similar study design: (a) cohorts of pregnant women were recruited prospectively; (b) vaginal swabs were collected during the pregnancies; (c) birth outcomes (e.g., PTB) were recorded; (d) 16S rRNA gene sequencing was performed on a subset of women selected to meet pre-specified inclusion criteria and a target PTB to term birth (TB) ratio. However, these studies reported varied and sometimes inconsistent associations between the vaginal microbiome and PTB. For example,Romero et al.(2014) found that vaginal microbial composition was not different in PTBs and TBs in a cohort of 90 pregnant women (88% African American; PTBs <34 weeks).DiGiulio et al.(2015) reported that lower Lactobacillus and higher *Gardnerella* abundances in the vaginal microbiome were associated with a higher risk of PTB (55%+ white; PTBs <37 weeks).Callahan et al.(2017) replicated these findings in a study cohort drawn from the same population as DiGiulio et al.(2015), but not in a different cohort with a prior history of PTB (82% African American; PTBs <37 weeks).Kindinger et al.(2017) found a lack of *Lactobacillus crispatus* and *Lactobacillus iners* dominance were risk factors for PTB in a cohort of UK women (65% white; PTBs <34 weeks). Fettweis et al.(2019) reported lower *L. crispatus* abundance was a risk factor for PTB in a predominantly black cohort of women (75% African American; PTBs <37 weeks). These studies employed a variety of different sequencing methodologies, including targeting different regions of the 16S rRNA gene, and varied in how they included and reported spontaneous versus indicated preterm births.

The substantial heterogeneities between previous studies almost certainly contribute to the variation in reported associations between the vaginal microbiome and PTB. These studies were highly heterogeneous in (1) the population being sampled (e.g., maternal race, BMI and age), (2) study size, (3) technical choices in microbiome profiling methods (e.g., DNA extraction method, 16S rRNA gene hypervariable region) and (4) definitions of term birth (TB) and PTB (e.g., spontaneous and indicated PTB, early and late PTB). Black women have a higher risk of PTB and BV compared to white women (Schaaf et al.,2013), and these study cohorts have very different racial compositions. Advanced maternal age is considered a risk factor of PTB, yet it is rarely accounted for in studies investigating the vaginal microbiome in pregnancy (Waldenström et al.,2014). Standard primers for the V4 hypervariable region of the 16S rRNA gene have higher sensitivity to the important *Gardnerella* genus compared with common V1-V3 primers (Frank et al.,2008;Mizrahi-Man et al.,2013). The mechanisms of spontaneous and indicated PTB might be different as spontaneous PTB is usually related to preterm rupture of membranes (PPROM) or cervical dilation while indicated PTB is related to induced or cesarean section labor due to obstetrical complications (Stout et al.,2014). Similarly, early PTB (< 32), moderate PTB (>= 32, < 34), and late PTB (>= 34, < 37) might be related to different causative factors, with an infectious etiology more commonly associated with early PTB (Goldenberg and Culhane, 2003). Lack of power also contributes to inconsistencies in reported associations. Many of these studies have small sample sizes — 12 cohorts included less than 50 women that experienced PTB, and 7 cohorts had an overall sample size of less than 100. This lack of power is exacerbated by the typically untargeted analyses that must account for the tens to thousands of taxa being simultaneously investigated.

In a meta-analysis, the results from multiple studies regarding a common biological question are synthesized to achieve greater power and generalizability of the conclusions. For example, two meta-analyses of the gut microbiome and colorectal cancer (CRC) performed integrated analyses of multiple metagenomic CRC datasets to reveal consistent associations between the gut microbiome and CRC and to better understand the reproducibility of such associations across studies (Thomas et al.,2019;Wirbel et al.,2019). Meta-analysis can help identify factors that cause inconsistencies between studies, aggregate signals across studies to improve power, and point the way towards improvements in future study design and analysis.Haque et al.(2017) pooled 4 vaginal microbiome datasets to understand the temporal differences between the vaginal microbiome communities and found the diversity measures are significantly different between vaginal microbiomes sampled from women with term and preterm outcomes. However, they did not look into the role of specific genera or species.Kosti et al.(2020) aggregated 5 longitudinal vaginal microbiome datasets with batch correction and reported several microbial genera as associated with PTB. However, they did not explore the heterogeneity cross-study and did not perform predictive analyses. Gudnadottir et al.(2022) performed a network-based meta-analysis of 17 longitudinal vaginal microbiome datasets using community state types (CSTs). Their results supported the predictivity of preterm birth using the vaginal microbiome but did not build any prediction models and CSTs reduce the description of the vaginal microbiome to the identity of its most abundant member.

In this study, we performed a meta-analysis of 12 prospective case-control PTB datasets obtained by using 16S rRNA gene sequencing to measure the vaginal microbiome during pregnancy. All together, these 12 datasets included 1926 pregnant women, 568 of whom went on to deliver preterm. After re-processing the raw sequencing data using a consistent bioinformatics pipeline, we used a machine learning approach to investigate the predictability of PTB from the composition of the vaginal microbiome in each study (Pasolli et al.,2016;Topçuoğlu et al.,2020). We evaluated cross-dataset reproducibility of PTB predictions from the vaginal microbiome, and investigated PTB and study-specific factors that affected prediction accuracy. We explored the association between specific microbial taxa and PTB within and across studies. Finally, we synthesized these results into specific recommendations and cautions applicable to future study of the vaginal microbiome in PTB, and perhaps to the study of microbiomes in health and disease more broadly.

## Results

### Published studies of the vaginal microbiome in term and preterm births are highly heterogeneous

We searched the published literature for studies between 2014 and 2020 that used high-throughput 16S rRNA gene sequencing to characterize the vaginal microbiome during pregnancy in term and preterm births. We identified 15 such studies, all of which used some variation of a nested case-control study design drawn from larger cohorts of women who were prospectively enrolled and sampled during pregnancy. From these 15 studies, we identified 12 datasets from independent cohorts of women that were complete enough (raw sequencing data and sufficient metadata) for us to include in this meta-analysis (Table1; Methods). In total, these datasets include 6,891 vaginal microbiome samples from 2,023 pregnant women, 570 of whom had preterm births.

**Table 1.**
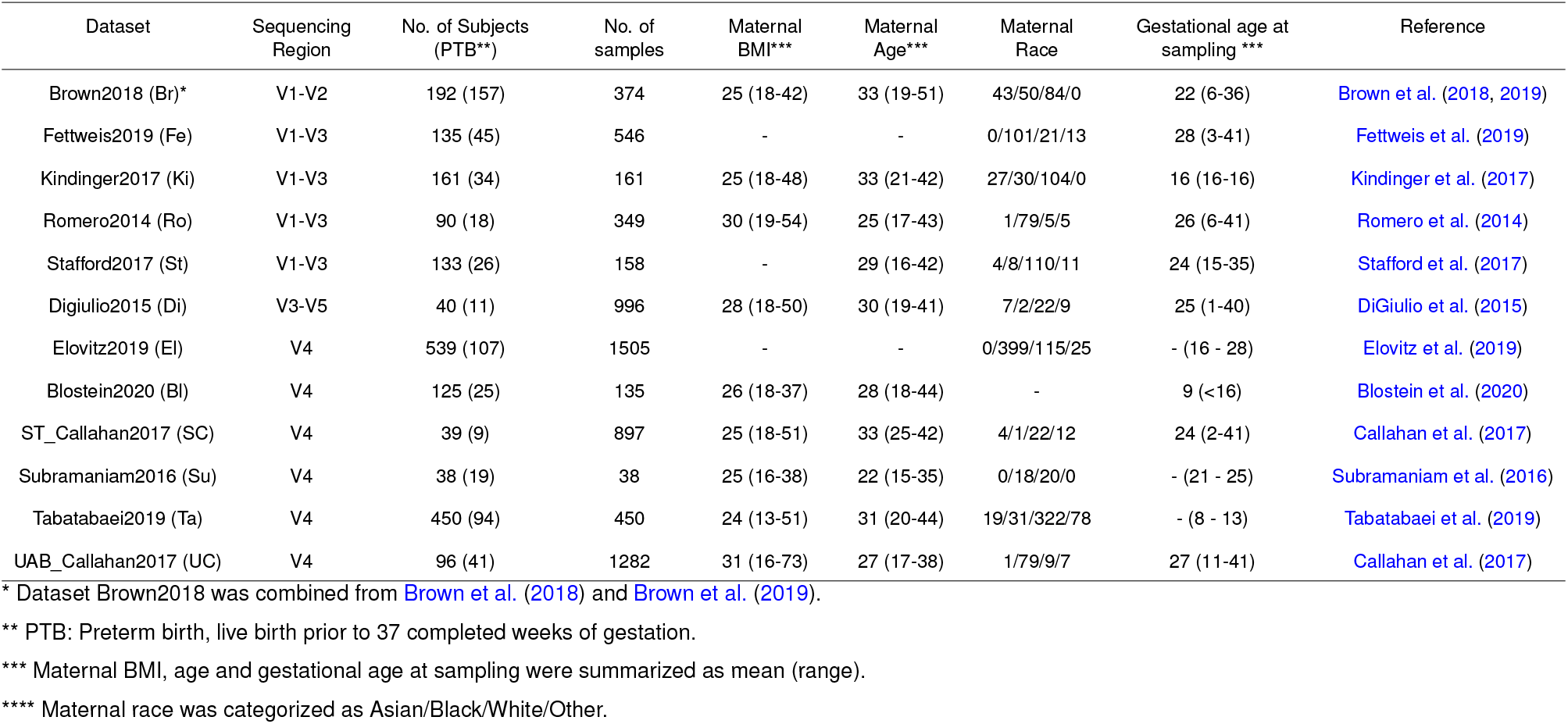
Characteristics of the 16S rRNA gene sequencing and study cohorts for each dataset included in this meta-analysis.

There was a large amount of heterogeneity in technical, clinical and cohort characteristics among these studies of the vaginal microbiome in term and preterm births. The number of subjects in the datasets we included in our meta-analysis varied from a low of 38 to a high of 539, and the percentage of subjects who went on to have preterm births varied from a low of 20% to a high of 81%, respectively.In terms of gestational age at sampling, three datasets (Ki, Bl, and Ta) obtained samples only from first trimester (0 - 13 weeks) or early of second trimester (14 - 26 weeks). Su datasets obtained samples only from late second trimester. All other eight datasets obtained samples across all trimesters. Furthermore, seven datasets are from longitudinal studies, with averages from 2.1 to 25 samples per subject. Furthermore, six datasets have subjects that included samples from all three trimesters. The number of subjects sampled in all trimesters varied from 3 to 45, and the number of those subjects who experienced PTB varied from 1 to 12 (Supplementary FigureS1; Supplementary TableS1).The V1-V2 region of the 16S rRNA gene was sequenced in five of these datasets, and the V4 gene region in the other seven. Four datasets had most participants self-report their race as black and six datasets had a majority of participants self-report their race as white. Three datasets excluded late PTB (>= 34, < 37) or early TB (>= 37, < 39), while all other datasets included these two categories. Seven datasets only included spontaneous PTB and at least two other datasets included both spontaneous and indicated PTB. The distributions of gestational age at delivery and select population characteristics also varied substantially between datasets (Supplementary FiguresS2&S3).

### A limited set of core taxa make up a vast majority of the vaginal microbiome

We used a consistent bioinformatic protocol based on the DADA2 tool and the Silva reference database to generate tables of taxonomically-assigned amplicon sequence variants (ASVs) from the raw 16S sequencing data for each dataset (Supplementary TablesS2&S3; Methods). To work at the highest level of resolution possible, datasets were partitioned into the V1-V2 or V4 groups based on which region of the 16S gene was sequenced. Within each group, ASVs were truncated to the “intersection” region contained within all sequenced amplicons, allowing an ASV table containing all datasets in the group to be constructed. For analyses using all datasets combined, taxonomic profiles at the genus level (with special species-level discrimination performed within the Lactobacillus genus) were merged into a single table.

A small number of taxa comprised a large majority of the vaginal microbiome in all studies included in our meta-analysis. In the V1-V2 group we found 42 “common” ASVs that were present in all datasets, and 157 “common” ASVs in the V4 group (Supplementary TableS4). These common ASVs constituted a large majority of the vaginal microbiome in every dataset in both the V1-V2 and V4 groups (71.3% - 94.8% of total reads). Another frequent strategy that is used to select a set of cross-dataset taxonomic features is to consider all taxa that appear above some abundance threshold in any dataset. Here we defined “top” ASVs as those that had a relative abundance larger than 0.1% in any dataset. We found 172 top ASVs and 159 top ASVs in the V4 group and the V1-V2 group, respectively. This still modest number of ASVs constituted an overwhelming majority of the vaginal microbiome in every dataset (90.4% - 98.0% total reads; 349 - 5479 ASVs). Finally, we also selected a set of “core” genera from the all-study table (both V1-V2 and V4 studies classified at the genus level, with *Lactobacillus* discriminated at the species level) using a hybrid filtering strategy that kept all genera present in at least 0.1% abundance and 10% prevalence in at least 5 datasets. Just 25 “core” genus-level taxa made up 88.4% - 97.1% of the total reads in every dataset (Figure1; Supplementary FiguresS4&S5). Five genera had particularly high average relative abundance (> 0.05) and prevalence (> 40%): *L. iners* (0.35, 87%), *L. crispatus* (0.29, 78%), *L. jensenii* (0.06, 57%), *L. gasseri* (0.057, 46%), and *Gardnerella* (0.054, 56%).

**Figure 1.**
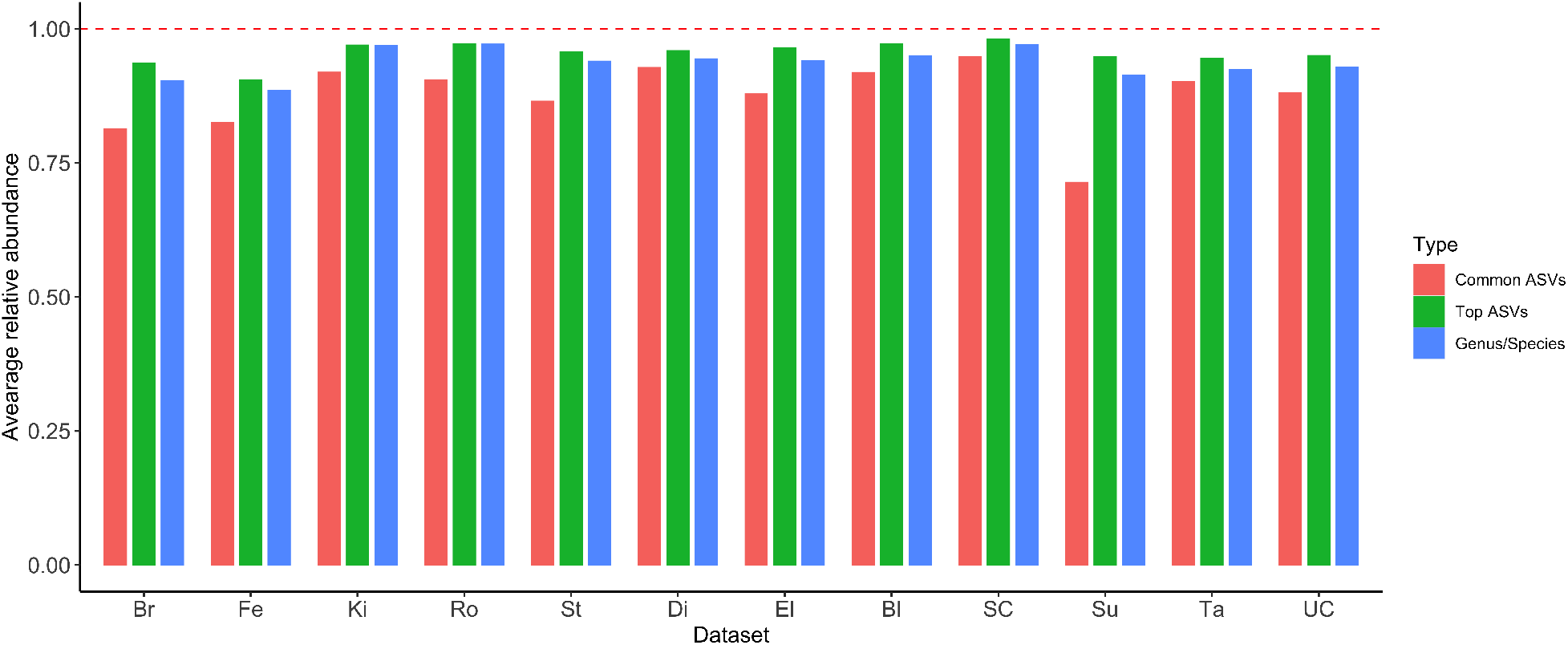
The average proportion of all sequencing reads in each dataset derived from the set of common ASVs (found in every dataset), top ASVs (proportion larger than 0.1% in any dataset) and core genus-level taxonomic features (abundance > 0.1% and prevalence > 10% for at least 5 datasets). Note that common and top ASV features were determined within the V1V2 and V4 dataset groups independently, as non-overlapping ASVs are not directly comparable (Methods).

### The predictivity of preterm birth from the vaginal microbiome is low

We employed a machine learning (ML) approach to assess the predictability of preterm birth outcomes from the genus-level composition of the vaginal microbiome. In order to assess the generalizability of ML predictions, we performed three types of ML analyses – intra-dataset analyses in which the ML model is trained and tested within a single dataset, cross-dataset analyses in which the ML model is trained on one dataset and tested on another, and leave-one-dataset-out (LODO) analyses in which 11 of 12 datasets are pooled together for training and testing is performed on the left-out dataset (Figure2A; Methods). Based on our evaluation of overall performance (Supplementary FiguresS6&S7; Supplementary Methods) and precedent in the microbiome field, we used the random forest classifier and either proportions or centered log-ratio (CLR) transformed abundances as our ML features. We used the area under the receiver operating characteristic curve (AUC) as our primary measure of ML prediction accuracy.

**Figure 2.**
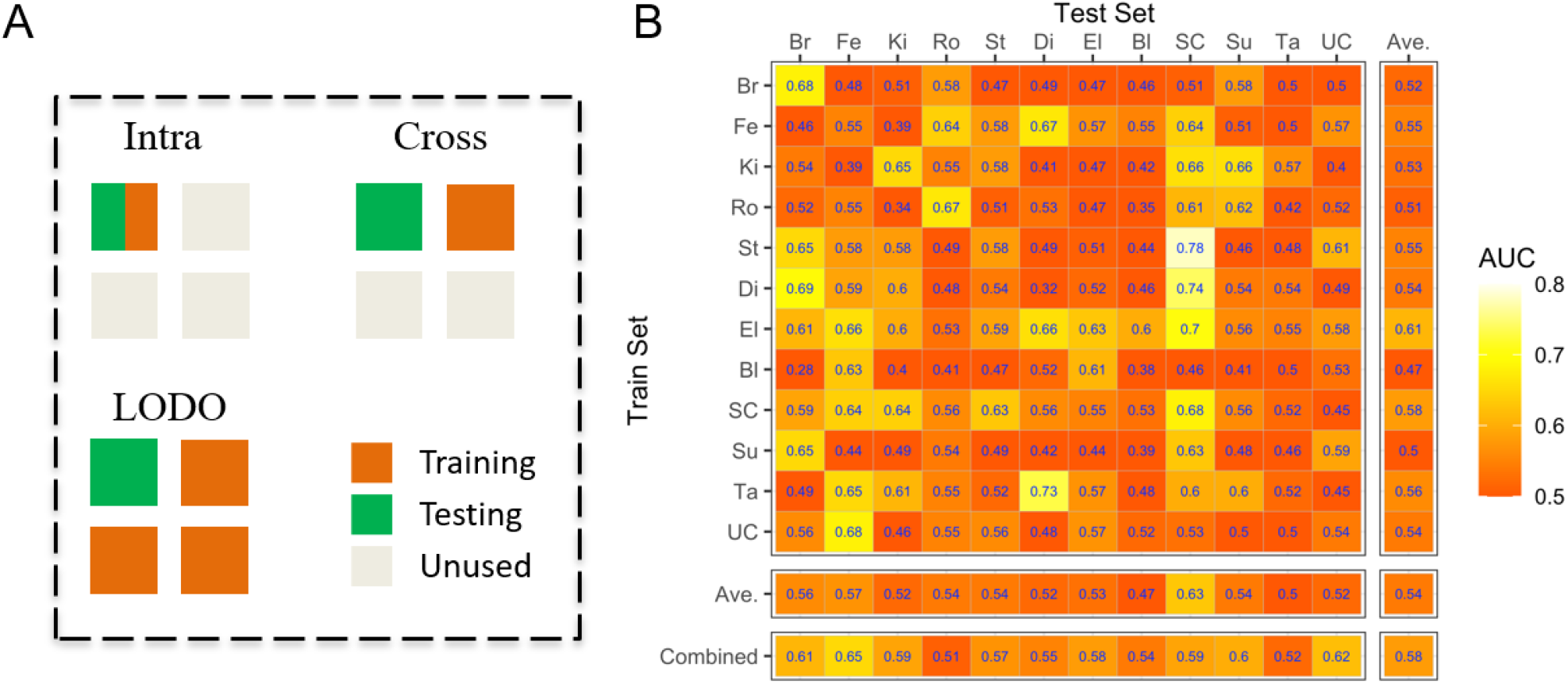
(A) A schematic of different analytical strategies using machine learning. Each square represents a different dataset, and squares are colored by how they are used to train or test the ML model. (B) The prediction accuracy, as measured by the AUC, for random forest ML models trained on the vaginal microbiome profiles (genus-level proportions) in one dataset (rows) and tested in the same or a different dataset (columns). “Ave.” indicates the average AUC of each row (same training dataset) or each column (same testing dataset).

The predictivity of PTB from the vaginal microbiome varied substantially across studies. Intra-dataset AUCs ranged from 0.32 (no predictivity) to 0.68 (moderate predictivity) across the datasets we considered (Figure2B, values in the major diagonal). We did not discern an obvious pattern amongst the higher or lower-predictivity datasets. There was no clear increase in AUC with study size: low intra-study AUC (0.52) was obtained in the relatively well-powered Ta study (n=450) while the highest intra-study AUC was in the relatively low power SC study (n=39). The three studies (Br, Ro, and SC) with the highest AUCs (0.67-0.68) include cohorts with predominantly white and predominantly black racial backgrounds, cohorts from California, Michigan and the United Kingdom, and variously considered both all-cause and only spontaneous preterm births.

The predictivity of PTB by ML models trained on data from a different dataset was generally low. In the cross-dataset analyses – in which the ML model is trained on one dataset and then used to predict on a different dataset – only 22% of training/testing dataset pairs yielded AUCs larger than 0.6, and just 3% had AUCs larger than 0.7. There was some indication that certain datasets were easier, or harder, to predict PTB in than others (columns of Figure2B). AUCs for predictions in the SC dataset were greater than 0.6 for ML models trained on most other datasets, while the AUCs for predictions in the Ta study never exceeded 0.57 for any training dataset. The leave-one-dataset-out (LODO) analysis yielded slight increases in AUC in 10 out of 12 datasets compared to the average cross-dataset AUC. While similar in direction to the previous results in the meta-analysis of the gut microbiome in Thomas et al. (2019) and Wirbel et al.(2019), the magnitude of the increase in prediction accuracy obtained by pooling datasets together for ML training was much smaller than observed in those studies.

### Earlier Preterm Birth Is More Predictable Than Late Preterm Birth

Preterm birth is a syndrome with multiple causes, the relative importance of which may vary between different populations, and between different sub-categories of preterm birth. One important subdivision of preterm birth is based on gestational age at delivery, with morbidity and mortality increasing sharply with earlier preterm births. Here, we used information available from each study about gestational age at delivery to define three PTB subgroups: (1) early preterm births (< 32 weeks); (2) early or moderate preterm births (< 34 weeks); and (3) late preterm births (>= 34 and < 37 weeks). Seven datasets had sufficient numbers of PTBs in each subgroup (8+) to include in our analysis. We employed intra-dataset and cross-dataset ML approaches to compare the predictivity of early, moderate and late preterm births, with the control group set to full-term births (>= 39 weeks). To completely remove any potential effect of preterm birth sample size from the results, within each study we resampled the number of women in each PTB subgroup to the smallest number of women among all subgroups (Methods). Unfortunately, due to both a lack of available data in some studies and the exclusion of indicated preterm births in other studies, we were unable to perform a similar analysis comparing indicated and spontaneous preterm births.

Earlier preterm births were much easier to predict from the composition of the vaginal microbiome than were later preterm births. The accuracy for predicting late PTB was low to moderate in all intra-dataset and cross-dataset ML analyses (all AUC values <= 0.65, Figure3C). In contrast, the accuracy for predicting early PTB was acceptable to good in most datasets (21 AUC values >= 0.65, 7 AUC values > 0.75, Figure3C). Classification accuracy for early-to-moderate PTB was intermediate, as expected. In most datasets the intra-dataset and LODO accuracy for predicting early PTB were substantially better than late PTB (Figure3A and 3B), but the Fe dataset was an exception where classifier performance remained poor (AUC < 0.6) for all categories of PTB. Similar results were observed whether using proportions or CLR-transformed abundances as the features in the analysis (Supplementary Figure S8).

**Figure 3.**
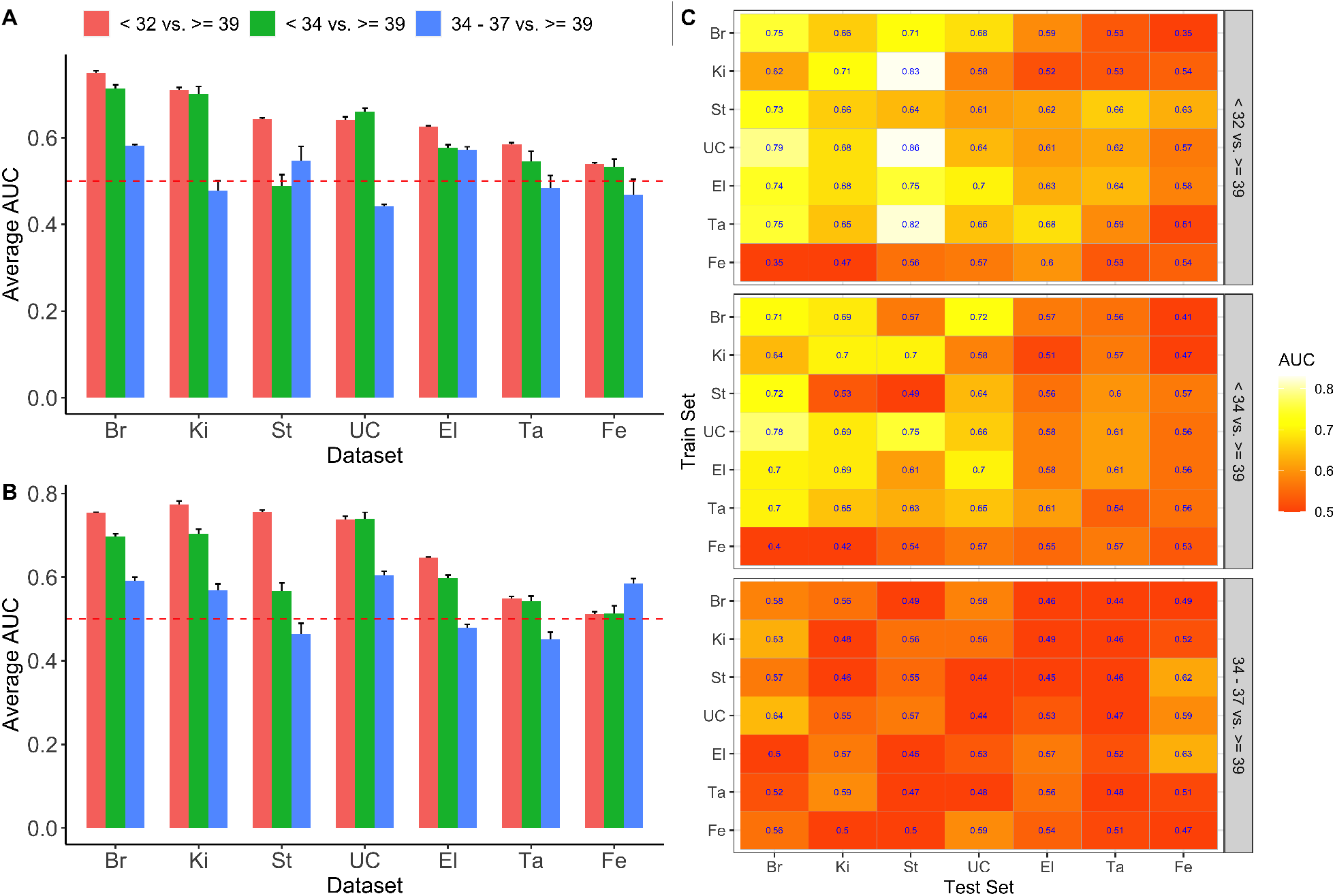
Assessment of prediction performance for different preterm birth groups using intra-dataset analysis (A), LODO analysis (B) and cross-dataset analysis (C) using CLR-transformed data. A resample procedure is used to ensure each preterm birth group has the same sample size. The experiment is repeated 10 times and the average AUC and/or standard error are calculated. For each heatmap, diagonal AUC values are from intra-analysis, and off diagonal values are from cross-analysis.

### More Resolved Taxonomic Features Inconsistently Improved the Predictivity of PTB

We compared the prediction accuracy of our random forest ML models trained on the proportions of taxonomic features at different levels of resolution, from ASV up to Phylum (Table2). In intra-dataset analysis, ASV level features had the overall best performance for the V1-V2 group, and there was an overall trend of decreasing AUC with increasingly broad taxonomic features. However, this trend was not evident in the V4 group (Table2). In both the V1-V2 and V4 groups of datasets, the AUC measured in the cross-dataset and LODO analyses showed no clear trend with the breadth of the taxonomic features considered. Given the previous observation that earlier preterm birth is more predictable, we further investigated the prediction accuracy at different levels of feature resolution using only early PTB (< 32 weeks) and late-term birth (>= 39 weeks). In the V1-V2 group, we observed the same overall trend of decreasing AUC with increasingly broad taxonomic features consistently in intra-dataset, cross-dataset, and LODO analysis. However, we again did not see a similar trend in the V4 group of datasets (Supplementary Table S5).

### The Importance of Microbial Taxa to Machine Learning Models Varies Across Datasets

We further investigated the ML models by computing the importance of genus-level microbial features using SHAP (SHapley Additive exPlanations) values (Lundberg and Lee,2017). Figure4 shows the feature ranking for each study for random forest models trained on proportional data. Averaged across all datasets, *L. crispatus* is the taxa that contributes most to the machine learning predictions, followed by *Prevotella* and *L. iners*. However, there is substantial variation in the importance of most taxa in ML models trained on different datasets. Consider, for example, *Finegoldia*. There are three datasets for which *Finegoldia* is among the three most important taxa and two studies for which it is among the three least important taxa. *Mycoplasma* is ranked as the most important taxa for the Bl dataset and the least important taxa for the Ta dataset. Further inconsistencies can be seen by examining the SHAP summary plot for each dataset (Supplementary FigureS9). For most studies for which *Prevotella* is among the most important features, a high relative abundance of *Prevotella* is associated with preterm birth (see the Br, Ki, and St datasets). However, a high relative abundance of *Prevotella* is associated with term birth in the Bl dataset.

**Figure 4.**
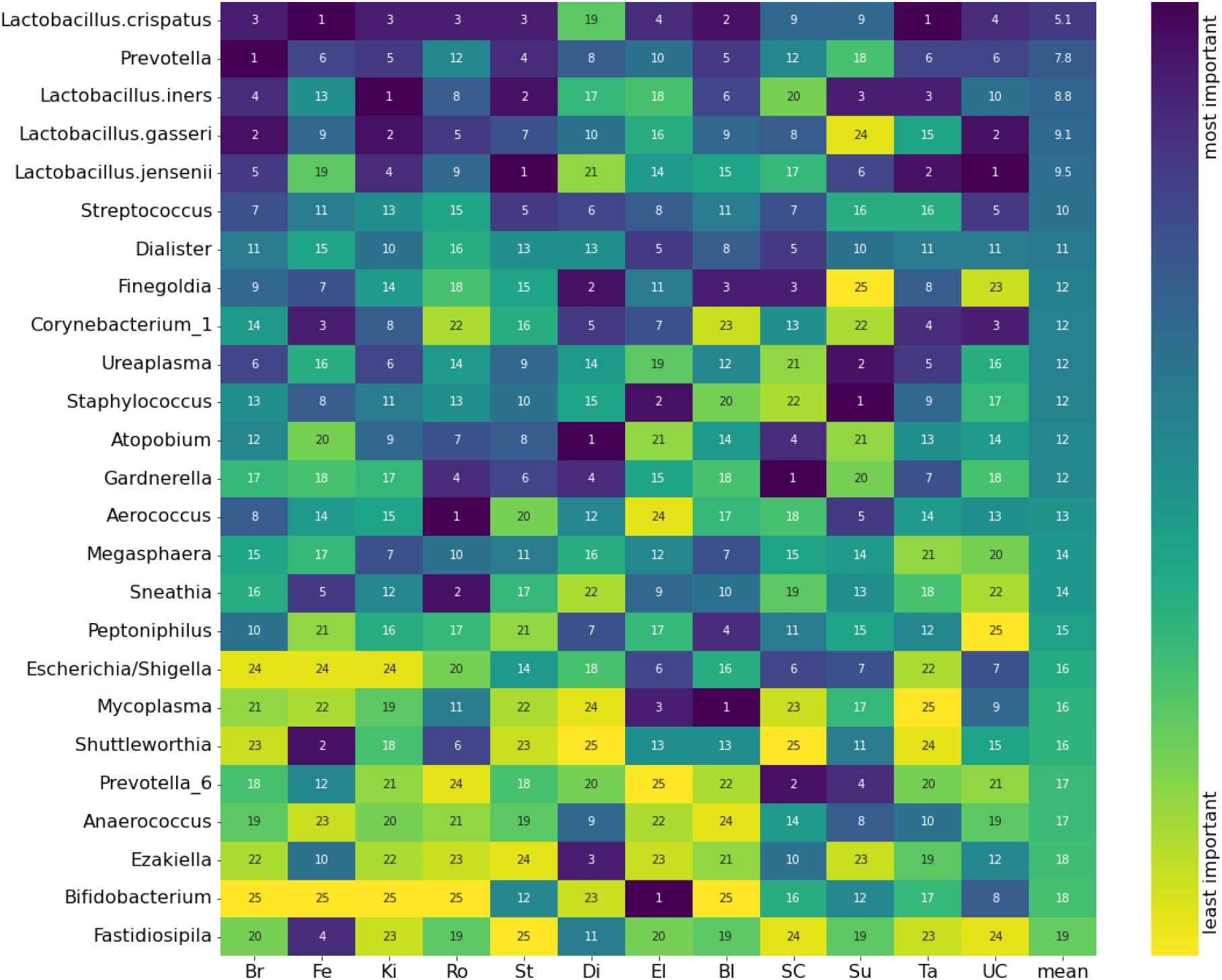
The feature importance ranking for genus-level taxonomic features (rows) in random forest ML models trained in different datasets (columns). Feature importance was quantified as the absolute SHAP value. Genera are ordered by their mean importance rank across all datasets.

The varying importance of taxa in different datasets contributes to the lower prediction accuracy for PTB in the cross-dataset and leave-one-dataset-out (LODO) analyses. As an example, *Aerococcus* was the most important feature for the Ro dataset but was unimportant for all other datasets. Machine learning models trained on the Ro study heavily weight the abundance of *Aerococcus* in their predictions, even though *Aerococcus* is a poor predictor of preterm birth in other datasets. We trained ten random forests on the Ro dataset with and without including *Aerococcus* as a feature and made predictions on the other studies (Supplementary FigureS10). For most studies, the AUC was higher when excluding *Aerococcus* from Ro, with notable improvements for the Fe, Di, SC, and Su studies.

### Emerging consensus associations between microbial genera and PTB

At an individual dataset level, differential abundance (DA) analysis using the one-sided Wilcoxon rank-sum test found associations between bacterial genera and PTB that largely agreed with the results reported in the original papers (Supplementary FigureS11). This re-analysis confirms that for many genera, there is too much variation in the effect sizes and even direction of their association with PTB to draw robust conclusions from individual dataset analyses. This is unsurprising given the low power of many of these datasets. However, taxa with more consistent directions of effect did emerge from when considering all the individual dataset DA results. In particular, L. crispatus was negatively associated with preterm birth in 10/12 datasets and *Gardnerella* was positively associated with preterm birth in 11/12 datasets.

In order to increase power, we performed an all-dataset differential prevalence analysis that also accounted for maternal age, BMI and self-reported race. We created a prevalence (presence-absence) table at the genus level for each study by defining a genus as present in a sample if its proportion was greater than 0.1%. We fit a generalized linear mixed model (GLMM) to estimate the odds ratio between a genus is present versus absent with dataset-specific random effects separately for each genus (Figure5; Supplementary FigureS12; Methods). The presence of three *Lactobacillus* species - *L. crispatus, L. jensenii* and *L. gasseri* - were associated with reduced risk of PTB, while the presence of L. iners was associated with increased risk of PTB. Based on unadjusted p-value at a 0.05 level, we observed significant associations of L. iners (p = 0.043) and *L. crispatus* (0.007). The presence of most non-Lactobacillus genera (18 out of 21; Figure5A) was associated with a higher risk of PTB, consistent with a higher-diversity “bacterial-vaginosis-like” vaginal microbiome being associated with PTB. There were significant positive associations between PTB and the presence of *Gardnerella* (p = 0.002), *Shuttleworthia* (p = 0.02), *Prevotella* (p = 0.0002), *Megasphaera* (p = 0.0007), *Atopobium* (p = 0.0001), *Sneathia* (p = 0.003), *Streptococcus* (p = 0.04), Dialister (p = 0.03) and *Mycoplasma* (p = 0.008). We further investigated these associations in PTBs subdivided into early, moderate and late subgroups as previously described (See Methods). We found that the associations between PTB and *L. iners, L. crispatus*, and *Prevotella* were stronger, i.e., had larger effect sizes and were more statistically significant, in earlier PTB than in late PTB (Supplementary FigureS13).

**Figure 5.**
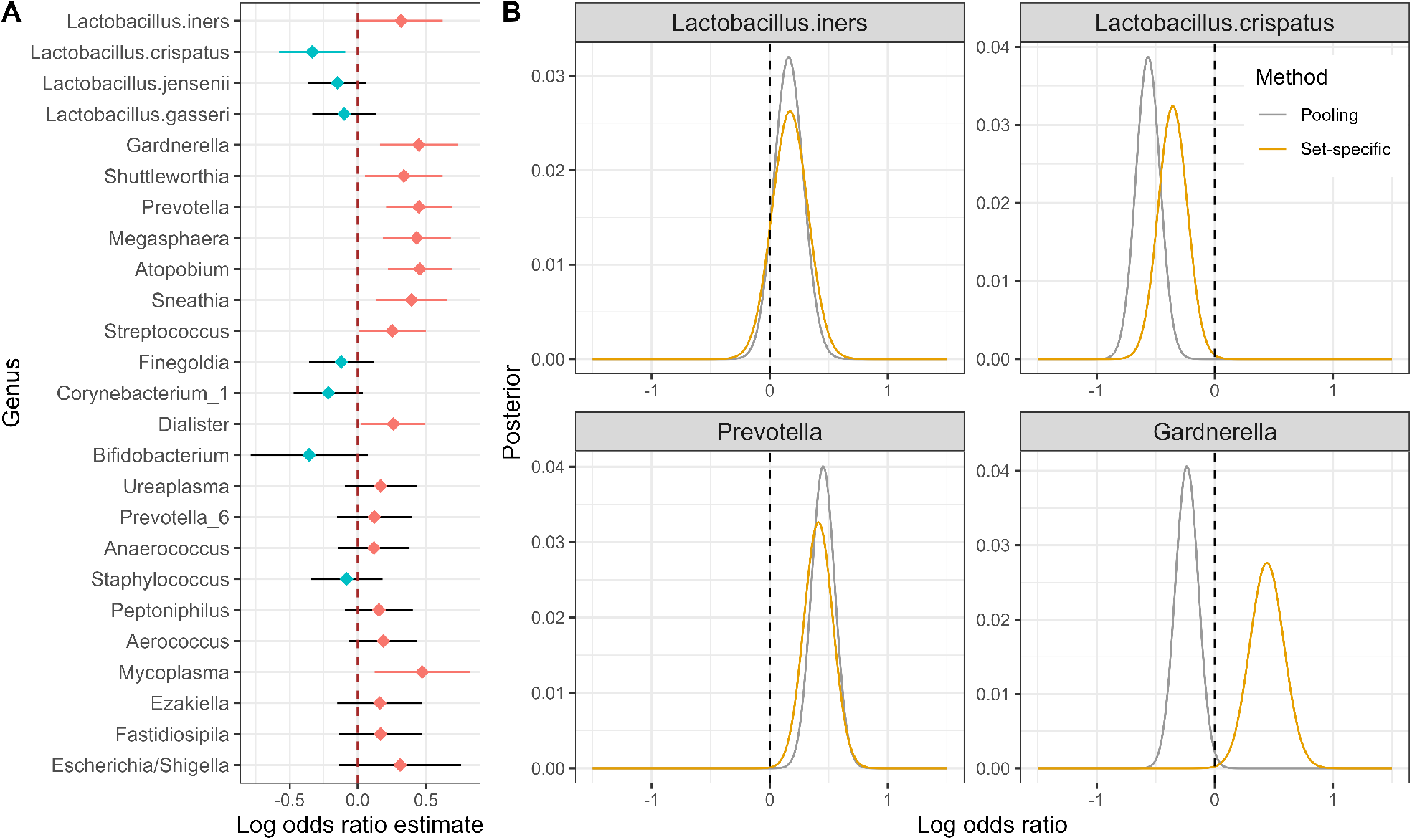
Cross-dataset differential abundance analysis. A: point estimates and 95% confidence interval of log odds ratio between a genus present and absent based on a relative abundance of 0.001 for each genus using a generalized linear mixed model. The model included all 12 datasets and no population characteristic covariates. Point estimates less than 0 are shown as blue points and greater than 0 as red points. Confident intervals less than 0 are shown as blue bars and greater than 0 as red bars. B: posterior distribution of log odds ratio using pooling and set-specific methods for four selected genus/species.

### Ignoring dataset-specific characteristics in microbiome analyses can cause false signals

We developed two related Bayesian analyses of the association between genus prevalence and PTB, one in which datasets were pooled together as exchangeable equals (Pooling) and another in which the baseline rate at which a taxon was detected was allowed to vary among datasets (Set-specific). More specifically, we performed two Bayesian analyses in which the log odds of a genus to be present given preterm birth and the odds ratio of PTB relative to TB follow prior distributions. We calculated the posterior distribution of the odds ratio between PTB and TB using two methods: (1) Pooling, in which all datasets were combined as a large dataset; and (2) Set-specific, in which the posterior distribution of the association between genus prevalence and preterm birth was sequentially updated by application to each dataset while allowing for a dataset-specific baseline rate at which that genus was detected. Consistent with the GLMM results reported above, in the set-specific method, 18 genera had a maximum a posteriori (MAP; mode of the posterior distribution) value of their odds larger than 0 (i.e., a positive association between their presence and PTB) and seven genera had MAP smaller than 0 (Supplementary FigureS14). Meaningful differences emerged when accounting for dataset-specific detection rates in some genera. For example, the set-specific method estimated a significant and positive association between the presence of *Gardnerella* and PTB, while the pooling method estimated a significant and negative association (Figure5B). Further inspection of this result revealed different detection rates of *Gardnerella* by most of the V1-V2 and the V4 studies. In Supplementary FigureS4B, for example, we observed that the prevalence of *Gardnerella* at three V1-V2 studies (Br: 10%, Ki: 7%, St: 25%) are much lower than in all V4 studies. This result suggests that accounting for dataset-specific detection rates might be important when aggregating results across microbiome studies.

## Discussion

The identification of robust associations between host-associated microbiomes and health outcomes remains an elusive goal in many areas of microbiome research, as highlighted by many examples of specific associations that did not reproduce across studies (Callahan et al.,2017;Huybrechts et al.,2020;Wirbel et al.,2019). There are several potential reasons for this. Most microbiome studies to date have been underpowered when considering the substantial temporal and inter-individual variability of host-associated microbial communities. Rapid progress in the laboratory and computational methods used to study microbiomes means that any two microbiome studies likely used measurement protocols different enough to make their results quantitatively incomparable (Martin,2019; McLaren et al.,2019;Tierney et al.,2022). The interaction between microbiomes and the host is mediated by poorly understood, and hence largely unrecorded, environmental and individual factors. And myriad other challenges that are generic beyond microbiome studies, such as differences between study populations and the criteria used to define cases and controls. With these challenges in mind, we used a machine-learning and meta-analysis approach to study the relationship between the vaginal microbiome and preterm birth across 12 independent datasets consisting of taxonomic profiles obtained by 16S rRNA gene sequencing of vaginal swabs obtained during gestations that resulted in term and preterm births. Overall, this analysis revealed substantial heterogeneity in the relationship between vaginal microbiome measurements and preterm birth outcomes from different studies. Yet, generalizable results and lessons also emerged, perhaps most importantly the higher predictivity of the vaginal microbiome for earlier preterm births.

Earlier preterm births (< 32 weeks, < 34 weeks) were more predictable from the composition of the vaginal microbiome than were late preterm births (34-37 weeks). This pattern was observed across most of the seven datasets included in our analysis of PTB sub-categories. It was observed both in ML models that were trained and tested in the same dataset and in ML models that were trained in one dataset and tested in another. The two datasets (Ta and Fe) in which this pattern was not evident were also the two datasets in which PTB was the least predictable overall. A strength of this analysis is the strong control of between-study differences: Comparisons are being made between early and late preterm births within a study, and the number of preterm births in each category per study is held constant. We believe these results support the prioritization of earlier preterm birth (< 34 weeks, or even earlier) in future studies of the relationship between the vaginal microbiome and PTB. Prioritization of earlier preterm births is consistent with their much higher morbidity and mortality. It is also supported by the arbitrariness of the 37-week cutoff, which can result in weak differences between term births (37 - 40 weeks) and the late preterm births (i.e., 34 - 37 weeks) that predominate in study cohorts targeting all preterm births.

Our meta-analysis of these 12 independent vaginal microbiome datasets increases the credibility of reported associations between *L. crispatus* and reduced risk of preterm birth, reported associations between *Gardnerella* and *Prevotella* and increased risk of preterm birth, and the different roles played by L. iners compared to other vaginal *Lactobacilli*. The most consistent finding across individual datasets was the negative association between the relative abundance of *L. crispatus* and preterm birth: 10 out of 12 datasets showed the same direction of effect. In 6 of these, the association had a raw p-value < 0.05. In our machine learning models, *L. crispatus* had the highest average importance across all studies for predicting PTB. When considering all datasets together, the association between the presence of *L. crispatus* and reduced risk of preterm birth was highly significant (p = 0.007) and was stronger for earlier preterm births. The association between *L. iners* and preterm birth was different from the other vaginal *Lactobacilli*: *L. iners* was associated with increased PTB risk in most individual studies, across all studies considered together, and more significantly so in earlier preterm births. Although the presence of several genera was associated with a higher risk of preterm birth when considering all studies together, *Gardnerella* was the genus most consistently associated with a higher risk of preterm birth at the individual-dataset level. *Prevotella* was the second most important taxa on average across our machine learning models. When all studies were considered together it had the second most significant association with PTB and even stronger effect size and statistical significance in earlier preterm birth.

Two example taxa, *Aerococcus* and *Gardnerella*, demonstrate the important ways that differences in taxon-specific detection rates across studies can alter measured associations between the microbiome and preterm birth. In the cross-dataset meta-analysis, *Aerococcus* comprised a small to vanishing fraction of the vaginal microbiome and the presence of *Aerococcus* was marginally associated with higher PTB risk. In contrast, in the Ro dataset, *Aerococcus* was significantly associated with decreased PTB risk and was detected at a significantly higher baseline rate. ML models trained on Ro have *Aerococcus* as the most important genus for predicting PTB, whereas *Aerococcus* has little to no importance in models trained on other datasets. The importance of *Aerococcus* in Ro-trained models reduces their prediction accuracy in other datasets; the cross-dataset accuracy of models trained on Ro is higher when the *Aerococcus* feature is removed prior to training. We do not know what is driving this significant difference in the detection rate of *Aerococcus*, it could reflect real differences between the populations studied in Ro versus other studies, or it could reflect methodological differences such as a higher detection efficiency for *Aerococcus* of the primer mixture used in the Ro study. In another example, the taxon *Gardnerella*, it has been known for some time that common V1 primers used in several studies here do not effectively amplify *Gardnerella* or the related *Bifidobacterium* (Frank et al.,2008;Romero et al.,2014). Consistent with this, we observed much lower proportions of *Gardnerella* in the V1-V2 studies, especially the Br, Ki, and St studies, that did not supplement their primer mixtures to detect *Gardnerella* better. Left unaddressed, this interfered with the cross-study estimation of the association between *Gardnerella* presence and preterm birth. Naively pooling the samples from all studies together led to the estimation of a negative association between *Gardnerella* and preterm birth. However, when our modeling incorporated study-specific differences in detection rates, *Gardnerella* was found to be significantly positively associated with preterm birth, in line with most reports in the literature.

The complex heterogeneities between different datasets significantly impede obtaining robustly generalizable results. The relative paucity of available subject metadata did not allow for post-hoc control of many individual characteristics thought to modulate the vaginal microbiome, PTB risk, or both. Although maternal race, age, BMI, and gestational age at delivery were available from most datasets, several other important metadata were only available in some or a few datasets. For example, the definition of spontaneous vs. indicated PTB was only available for three datasets, while two datasets reported mixed spontaneous or indicated PTB. Prior history of preterm birth, which is a known risk factor for PTB, was only recorded for three datasets. Other complications such as pre-eclampsia and gestational diabetes, which are associated with a higher risk of PTB, were also underreported. Data on feminine hygiene practices such as douching was unavailable for several studies and has recently been reported to alter the relationship between the vaginal microbiome and preterm birth in white women (Nieves-Ramírez et al.,2021). It is therefore important that future studies capture and report comprehensive and detailed patient metadata that permit deeper analyses of potential confounding (Mirzayi et al.,2021).

When considering all-cause and all-type PTB, we found that the predictivity of PTB from the composition of the vaginal microbiome was low to modest. This should not be surprising. PTB is a syndrome with multiple causes, and it is highly unlikely that PTBs arising from different causal mechanisms, e.g., indicated PTB due to placenta previa versus spontaneous preterm labor due to intrauterine infection (Chan et al.,2022), will associate with similar patterns in the vaginal microbiome. Indeed it is likely that some etiologies of preterm birth will have no measurable relationship with the vaginal microbiome, and their inclusion in ML models would restrict the predictive accuracy. However, prediction of well-defined subtypes of PTB from the vaginal microbiome with moderate to high accuracy needed for clinical relevance may be achievable.

The cross-dataset and leave-one-dataset-out machine learning results highlight the importance of careful interpretation when evaluating the generalizability of machine learning classifiers trained on microbiome data and indicate that single-study examples of high AUC should be met with caution. Figure2B presents a classifier trained on the Br dataset that performs moderately well when tested on withheld test data from the same study (AUC = 0.68). However, it performs much worse when tested on data from other studies (average AUC = 0.52). This same behavior can be seen in previous microbiome meta-analyses that explored cross-dataset predictivity using machine learning (Thomas et al.,2019;Wirbel et al.,2019). However, such analyses can also highlight differences in the underlying microbiota-host interactions of different patient cohorts. The Br dataset was enriched for PTB cases preceded by preterm prelabor rupture of the fetal membranes (PPROM), which is often associated with an infectious etiology (Bennett et al.,2020). Thus, despite producing a highly accurate within-study classifier, the performance of this classifier may not be maintained in other cohorts where population characteristics (e.g., PPROM prevalence) differ substantially. Methodological differences in DNA extraction or sequencing may also restrict cross-study classification accuracy. Consistent with this, we found that combining datasets for training ML models (the LODO analyses) did not meaningfully improve cross-dataset prediction accuracy versus using a single dataset for training. This is different than the meaningful improvements in accuracy obtained by pooling studies together reported for predicting colorectal cancer from the gut microbiome in Thomas et al.(2019) and Wirbel et al.(2019). The reasons for this difference remain unclear but could arise from the higher heterogeneity of vaginal microbiome studies or even the PTB phenotype itself.

Finally, we make three suggestions for future studies of the vaginal microbiome and PTB studies: (1) earlier preterm birth should be prioritized, (2) the core genera discussed in this meta-analysis should be captured in future studies to reflect the community of the vaginal microbiome and (3) comprehensive subject metadata should be recorded, accounted for within data modeling and made available to the wider research community. Specifically, we recommend capturing and reporting maternal race, age, BMI, prior history of PTB, the use of interventions designed to prevent preterm birth, gestational age at delivery, gestational age at the time of sample collection, and whether PTB was spontaneous or indicated.

## Methods

### Data Collection and Availability

We included 12 datasets that at least have raw sequence data and minimal metadata, including subject/sample ID identifier and gestational age at sampling, that are available to us. Additional metadata were also collected if they are available, including maternal age, BMI, self-reported race, prior PTB, PPROM, PTB type (spontaneous or indicate), gestational complication and intervention. The raw sequence data and metadata were downloaded from the NCBI Sequence Read Archive, or obtained from the supplementary materials of original papers, or directly shared by authors. See Supplementary TableS1 for details.

### Bioinformatics

DADA2 (Callahan et al.,2016) was used to process the raw sequence data and infer the amplicon sequence variants (ASVs). The details of the DADA2 pipeline for each study can be found in Supplementary Table 2. To obtain comparable ASVs among datasets, we divided the datasets into two groups based on the region of the 16S gene that was sequenced (V1-V2 and V4) with five datasets in the V1-V2 group and seven datasets in the V4 group. Then we truncated the original ASVs separately for each group to a common V1-V2 or V4 region in three steps: (1) align the original ASVs to the SILVA reference database using the mothur software (Schloss et al.,2009); (2) identify the overlapping sequencing region common to all ASVs in the group using an alignment visualization tool (MSAviewer); (3) truncate the original ASVs and remove alignment gaps using the extractalign and degapseq commands. Furthermore, we assigned the taxonomy levels to each truncated ASV using DADA2. *Lactobacillus* species were assigned manually using BLAST against sequences from cultured *Lactobacillus* strains.

**Table 2.** Average AUC values for different taxa level features.

| Analysis | V1-V2 group |  |  |  |  |  | V4 group |  |  |  |  |  |
| --- | --- | --- | --- | --- | --- | --- | --- | --- | --- | --- | --- | --- |
|  | ASV | Genus | Family | Order | Class | Phylum | ASV | Genus | Family | Order | Class | Phylum |
| Intra | 0.65 | 0.63 | 0.60 | 0.57 | 0.58 | 0.55 | 0.58 | 0.57 | 0.59 | 0.56 | 0.52 | 0.54 |
| Cross | 0.53 | 0.54 | 0.57 | 0.56 | 0.56 | 0.57 | 0.57 | 0.56 | 0.56 | 0.55 | 0.53 | 0.55 |
| LODO | 0.56 | 0.57 | 0.57 | 0.58 | 0.60 | 0.61 | 0.60 | 0.62 | 0.61 | 0.60 | 0.56 | 0.59 |

### Data processing

Samples with total reads less than 100 were excluded in the analysis. The 25 core genera/species were obtained by the following steps: (1) assign the genus-level (species-level for *Lactobacillus*) name to “common” ASVs and “top” ASVs for both the V1-V2 and V4 groups; (2) match the genus level features between V1-V2 and V4 groups from both “common” and “top” sets; (3) filter the genera using the following criteria (i) at least 5 datasets have the average relative abundance > 0.1%; (ii) at least 5 datasets have the average prevalence > 10% (Supplementary FigureS4). For longitudinal datasets, we obtained the subject-level proportions of each genus by averaging the proportions from samples in the same subject.

### Machine Learning

A machine learning framework using random forest classifier was employed for intra-dataset, cross-dataset, and leave-one-dataset-out (LODO) analysis. Intra-dataset analysis was performed on a single dataset using 5-fold cross-validation. Cross-dataset analysis was performed based on a pair of two datasets: one dataset as a training set and the other as a testing set. LODO analysis was performed by combining all datasets as a training set except one hold-out dataset as a testing set (see Figure2A). Given the predicted and true results, the area under the receiver operating characteristic curve (AUC) is calculated using the ‘pROC’ R package. The AUC from intra-analysis was averaged among 20 repetitions. We performed ten repetitions of intra-dataset, cross-dataset and LODO analysis in the preterm birth subgroup analysis and calculated the average AUC. The random forest classifier was implemented using the ‘randomForest’ R package. We set the hyperparameter nTree = 1000, and tuned mtry using the ‘caret’ R package.

### Feature Importance

We used the SHAP value from the random forest classifier to determine the feature importance. SHAP values that are large in absolute value indicate that the corresponding feature was influential in the machine learning prediction for the given sample. Here positive SHAP values indicate that the feature value is associated with preterm birth whereas negative SHAP values indicate that the feature value is associated with term birth. We trained random forest models on each dataset using the scikit-learn implementation of a random forest classifier. For each dataset, we used 5-fold cross-validation with ten repetitions and calculated the SHAP values (Lundberg and Lee,2017) of the validation data using the Tree SHAP algorithm (Lundberg et al.,2020) as implemented in the SHAP package. The features were ranked according to the importance of each study by comparing the mean absolute SHAP values.

### Data Transformation

Due to the arbitrary sequencing read depth of each sample, we first converted count data obtained from the DADA2 pipeline to proportional abundance data by dividing the read depth of each sample. As the proportional abundance data does not account for the compositionality of the microbiome data, we further performed the centered log-ratio (CLR) transformation (with 10^−6^ pseudo-abundance) and compared the performance with proportional data in the ML framework and DA analysis. We found that data with CLR transformation have a similar prediction accuracy with the proportional abundance data using the ML model. The one-side Wilcoxon rank-sum test using CLR-transformed data or proportional data sometimes give an opposite direction of effect. For genera with low prevalence and low abundance, if CLR-transformed data is used to do analysis, their effects on preterm birth are mostly determined by the geometric mean of all genera, instead of their own abundance. However, if proportional data are used, their effects are only determined by their own abundance. In addition, we also included additive log-ratio (ALR), natural logarithm (log), and rank transformation methods in the ML framework comparison.

### Statistical Analysis

A one-sided Wilcoxon rank-sum test was used to perform the dataset-specific differential abundance analysis and implemented using the wilcox.test function in R. A generalized linear mixed model was used to do differential abundance analysis with a random effect for each study. Specifically, for each genus we fitted the following model:

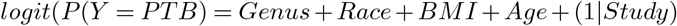

Genus=1 if a genus is present based on an abundance threshold of 0.001 and 0 otherwise. Race represents the maternal race; BMI represents the maternal BMI; Age represents the maternal age. The generalized linear mixed model was implemented using the glmer function in the ‘lme4’ R package. Due to missing maternal race, BMI, and age in some datasets and non-significance of these covariates except self-reported Black race vs. white race (p = 0.04), we also fitted the model without adjusting these covariates.

We further performed a Bayesian analysis by assuming that (1) the log odds of the presence of a genus given preterm birth follow a uniform prior distribution for each dataset; (2) the odds ratio between PTB and TB has the same underlying true distribution for each dataset. We use a uniform prior distribution for the odds ratio for the first dataset, then calculate the posterior distribution. Then we let the posterior distribution of the odds ratio from the first dataset be the prior distribution for the second dataset and update the posterior distribution. We repeated the process until the last dataset to obtain the final posterior distribution of the odds ratio. See the Supplementary Methods section for more details.

## Data Availability

ENA/SRA or dbGap with reference numbers summarized in the supplementary table S2. The processed data are available in the first author's github by request.

## Ethics Statement

The study was approved by the Institutional Review Board of North Carolina State University (Protocol Number 23575).

## Bibliography

Bennett, P. R., Brown, R. G., and MacIntyre, D. A. Vaginal microbiome in preterm rupture of membranes. Obstetrics and Gynecology Clinics, 47(4):503–521, 2020.

Blostein, F., Gelaye, B., Sanchez, S. E., Williams, M. A., and Foxman, B. Vaginal microbiome diversity and preterm birth: results of a nested case–control study in peru. Annals of epidemiology, 41:28–34, 2020.

Brown, R. G., Marchesi, J. R., Lee, Y. S., Smith, A., Lehne, B., Kindinger, L. M., Terzidou, V., Holmes, E., Nicholson, J. K., Bennett, P. R., et al. Vaginal dysbiosis increases risk of preterm fetal membrane rupture, neonatal sepsis and is exacerbated by erythromycin. BMC medicine, 16(1):1–15, 2018.

Brown, R. G., Al-Memar, M., Marchesi, J. R., Lee, Y. S., Smith, A., Chan, D., Lewis, H., Kindinger, L., Terzidou, V., Bourne, T., et al. Establishment of vaginal microbiota composition in early pregnancy and its association with subsequent preterm prelabor rupture of the fetal membranes. Translational Research, 207:30–43, 2019.

Callahan, B. J., McMurdie, P. J., Rosen, M. J., Han, A. W., Johnson, A. J. A., and Holmes, S. P. Dada2: High-resolution sample inference from illumina amplicon data. Nature methods, 13(7):581–583, 2016.

Callahan, B. J., DiGiulio, D. B., Goltsman, D. S. A., Sun, C. L., Costello, E. K., Jeganathan, P., Biggio, J. R., Wong, R. J., Druzin, M. L., Shaw, G. M., et al. Replication and refinement of a vaginal microbial signature of preterm birth in two racially distinct cohorts of us women. Proceedings of the National Academy of Sciences, 114(37):9966–9971, 2017.

Chan, D., Bennett, P. R., Lee, Y. S., Kundu, S., Teoh, T., Adan, M., Ahmed, S., Brown, R. G., David, A. L., Lewis, H. V., et al. Microbial-driven preterm labour involves crosstalk between the innate and adaptive immune response. Nature communications, 13(1):1–15, 2022.

Chawanpaiboon, S., Vogel, J. P., Moller, A.-B., Lumbiganon, P., Petzold, M., Hogan, D., Landoulsi, S., Jampathong, N., Kongwattanakul, K., Laopaiboon, M., et al. Global, regional, and national estimates of levels of preterm birth in 2014: a systematic review and modelling analysis. The Lancet Global Health, 7(1):e37–e46, 2019.

DiGiulio, D. B., Callahan, B. J., McMurdie, P. J., Costello, E. K., Lyell, D. J., Robaczewska, A., Sun, C. L., Goltsman, D. S., Wong, R. J., Shaw, G., et al. Temporal and spatial variation of the human microbiota during pregnancy. Proceedings of the National Academy of Sciences, 112(35):11060–11065, 2015.

Donders, G., Van Calsteren, K., Bellen, G., Reybrouck, R., Van den Bosch, T., Riphagen, I., and Van Lierde, S. Predictive value for preterm birth of abnormal vaginal flora, bacterial vaginosis and aerobic vaginitis during the first trimester of pregnancy. BJOG: An International Journal of Obstetrics & Gynaecology, 116(10):1315–1324, 2009.

Elovitz, M. A., Gajer, P., Riis, V., Brown, A. G., Humphrys, M. S., Holm, J. B., and Ravel, J. Cervicovaginal microbiota and local immune response modulate the risk of spontaneous preterm delivery. Nature communications, 10(1):1–8, 2019.

Ferrero, D. M., Larson, J., Jacobsson, B., Di Renzo, G. C., Norman, J. E., Martin Jr, J. N., D’alton, M., Castelazo, E., Howson, C. P., Sengpiel, V., et al. Cross-country individual participant analysis of 4.1 million singleton births in 5 countries with very high human development index confirms known associations but provides no biologic explanation for 2/3 of all preterm births. PloS one, 11(9):e0162506, 2016.

Fettweis, J. M., Serrano, M. G., Brooks, J. P., Edwards, D. J., Girerd, P. H., Parikh, H. I., Huang, B., Arodz, T. J., Edupuganti, L., Glascock, A. L., et al. The vaginal microbiome and preterm birth. Nature medicine, 25(6):1012–1021, 2019.

Frank, J. A., Reich, C. I., Sharma, S., Weisbaum, J. S., Wilson, B. A., and Olsen, G. J. Critical evaluation of two primers commonly used for amplification of bacterial 16s rrna genes. Applied and environmental microbiology, 74(8):2461–2470, 2008.

Goldenberg, R. L. and Culhane, J. F. Infection as a cause of preterm birth. Clinics in perinatology, 30(4):677–700, 2003.

Goldenberg, R. L., Culhane, J. F., Iams, J. D., and Romero, R. Epidemiology and causes of preterm birth. The lancet, 371(9606): 75–84, 2008.

Gudnadottir, U., Debelius, J. W., Du, J., Hugerth, L. W., Danielsson, H., Schuppe-Koistinen, I., Fransson, E., and Brusselaers, N. The vaginal microbiome and the risk of preterm birth: a systematic review and network meta-analysis. Scientific reports, 12 (1):1–8, 2022.

Haque, M. M., Merchant, M., Kumar, P. N., Dutta, A., and Mande, S. S. First-trimester vaginal microbiome diversity: A potential indicator of preterm delivery risk. Scientific reports, 7(1):1–10, 2017.

Huybrechts, I., Zouiouich, S., Loobuyck, A., Vandenbulcke, Z., Vogtmann, E., Pisanu, S., Iguacel, I., Scalbert, A., Indave, I., Smelov, V., et al. The human microbiome in relation to cancer risk: A systematic review of epidemiologic studiesthe human microbiome and cancer risk. Cancer Epidemiology, Biomarkers & Prevention, 29(10):1856–1868, 2020.

Kindinger, L. M., Bennett, P. R., Lee, Y. S., Marchesi, J. R., Smith, A., Cacciatore, S., Holmes, E., Nicholson, J. K., Teoh, T., and MacIntyre, D. A. The interaction between vaginal microbiota, cervical length, and vaginal progesterone treatment for preterm birth risk. Microbiome, 5(1):1–14, 2017.

Kosti, I., Lyalina, S., Pollard, K. S., Butte, A. J., and Sirota, M. Meta-analysis of vaginal microbiome data provides new insights into preterm birth. Frontiers in Microbiology, 11:476, 2020.

Lundberg, S. M. and Lee, S.-I. A unified approach to interpreting model predictions. Advances in neural information processing systems, 30, 2017.

Lundberg, S. M., Erion, G., Chen, H., DeGrave, A., Prutkin, J. M., Nair, B., Katz, R., Himmelfarb, J., Bansal, N., and Lee, S.-I. From local explanations to global understanding with explainable ai for trees. Nature machine intelligence, 2(1):56–67, 2020.

Manuck, T. A., Rice, M. M., Bailit, J. L., Grobman, W. A., Reddy, U. M., Wapner, R. J., Thorp, J. M., Caritis, S. N., Prasad, M., Tita, A. T., et al. Preterm neonatal morbidity and mortality by gestational age: a contemporary cohort. American journal of obstetrics and gynecology, 215(1):103–e1, 2016.

Martin, J. Reproducibility: the search for microbiome standards, 2019.

McLaren, M. R., Willis, A. D., and Callahan, B. J. Consistent and correctable bias in metagenomic sequencing experiments. Elife, 8:e46923, 2019.

Mirzayi, C., Renson, A., Zohra, F., Elsafoury, S., Geistlinger, L., Kasselman, L. J., Eckenrode, K., van de Wijgert, J., Loughman, A., Marques, F. Z., et al. Reporting guidelines for human microbiome research: the storms checklist. Nature medicine, 27(11): 1885–1892, 2021.

Mizrahi-Man, O., Davenport, E. R., and Gilad, Y. Taxonomic classification of bacterial 16s rrna genes using short sequencing reads: evaluation of effective study designs. PloS one, 8(1):e53608, 2013.

Nieves-Ramírez, M., Partida-Rodríguez, O., Moran, P., Serrano-Vázquez, A., Pérez-Juárez, H., Pérez-Rodríguez, M., Arrieta, M., Ximénez-García, C., and Finlay, B. Cervical squamous intraepithelial lesions are associated with differences in the vaginal microbiota of mexican women. Microbiology Spectrum, 9(2):e00143–21, 2021.

Onderdonk, A. B., Delaney, M. L., and Fichorova, R. N. The human microbiome during bacterial vaginosis. Clinical microbiology reviews, 29(2):223–238, 2016.

Pasolli, E., Truong, D. T., Malik, F., Waldron, L., and Segata, N. Machine learning meta-analysis of large metagenomic datasets: tools and biological insights. PLoS computational biology, 12(7):e1004977, 2016.

Romero, R., Hassan, S. S., Gajer, P., Tarca, A. L., Fadrosh, D. W., Bieda, J., Chaemsaithong, P., Miranda, J., Chaiworapongsa, T., and Ravel, J. The vaginal microbiota of pregnant women who subsequently have spontaneous preterm labor and delivery and those with a normal delivery at term. Microbiome, 2(1):1–15, 2014.

Schaaf, J. M., Liem, S. M., Mol, B. W. J., Abu-Hanna, A., and Ravelli, A. C. Ethnic and racial disparities in the risk of preterm birth: a systematic review and meta-analysis. American journal of perinatology, 30(06):433–450, 2013.

Schloss, P. D., Westcott, S. L., Ryabin, T., Hall, J. R., Hartmann, M., Hollister, E. B., Lesniewski, R. A., Oakley, B. B., Parks, D. H., Robinson, C. J., et al. Introducing mothur: open-source, platform-independent, community-supported software for describing and comparing microbial communities. Applied and environmental microbiology, 75(23):7537–7541, 2009.

Stafford, G. P., Parker, J. L., Amabebe, E., Kistler, J., Reynolds, S., Stern, V., Paley, M., and Anumba, D. O. Spontaneous preterm birth is associated with differential expression of vaginal metabolites by lactobacilli-dominated microflora. Frontiers in physiology, 8:615, 2017.

Stout, M. J., Busam, R., Macones, G. A., and Tuuli, M. G. Spontaneous and indicated preterm birth subtypes: interobserver agreement and accuracy of classification. American journal of obstetrics and gynecology, 211(5):530–e1, 2014.

Subramaniam, A., Kumar, R., Cliver, S. P., Zhi, D., Szychowski, J. M., Abramovici, A., Biggio, J. R., Lefkowitz, E. J., Morrow, C., and Edwards, R. K. Vaginal microbiota in pregnancy: evaluation based on vaginal flora, birth outcome, and race. American journal of perinatology, 33(04):401–408, 2016.

Tabatabaei, N., Eren, A., Barreiro, L., Yotova, V., Dumaine, A., Allard, C., and Fraser, W. Vaginal microbiome in early pregnancy and subsequent risk of spontaneous preterm birth: a case–control study. BJOG: An International Journal of Obstetrics & Gynaecology, 126(3):349–358, 2019.

Thomas, A. M., Manghi, P., Asnicar, F., Pasolli, E., Armanini, F., Zolfo, M., Beghini, F., Manara, S., Karcher, N., Pozzi, C., et al. Metagenomic analysis of colorectal cancer datasets identifies cross-cohort microbial diagnostic signatures and a link with choline degradation. Nature medicine, 25(4):667–678, 2019.

Tierney, B. T., Tan, Y., Yang, Z., Shui, B., Walker, M. J., Kent, B. M., Kostic, A. D., and Patel, C. J. Systematically assessing microbiome–disease associations identifies drivers of inconsistency in metagenomic research. PLoS biology, 20(3):e3001556, 2022.

Topçuoğlu, B. D., Lesniak, N. A., Ruffin IV, M. T., Wiens, J., and Schloss, P. D. A framework for effective application of machine learning to microbiome-based classification problems. MBio, 11(3):e00434–20, 2020.

Waldenström, U., Aasheim, V., Nilsen, A. B. V., Rasmussen, S., Pettersson, H. J., and Shytt, E. Adverse pregnancy outcomes related to advanced maternal age compared with smoking and being overweight. Obstetrics & Gynecology, 123(1):104–112, 2014.

Wirbel, J., Pyl, P. T., Kartal, E., Zych, K., Kashani, A., Milanese, A., Fleck, J. S., Voigt, A. Y., Palleja, A., Ponnudurai, R., et al. Meta-analysis of fecal metagenomes reveals global microbial signatures that are specific for colorectal cancer. Nature medicine, 25(4):679–689, 2019.

