## Supplemental materials for "Meta-Analysis Reveals the Vaginal Microbiome is a Better Predictor of Earlier Than Later Preterm Birth"

### Supplementary Figures

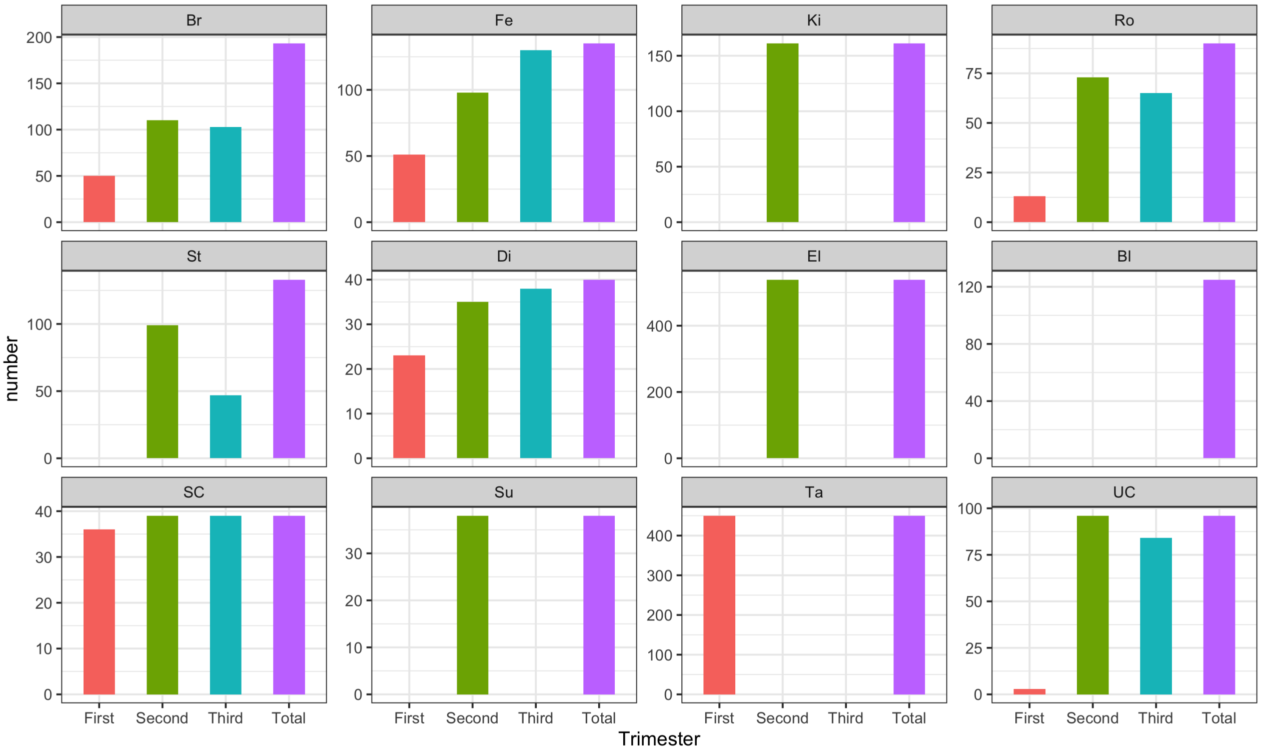


**Figure S1.** Number of subjects in each trimester during the pregnancy. First trimester: 0 to 13 weeks; second trimester: 14 to 26 weeks; third trimester: 27 to 40 weeks.

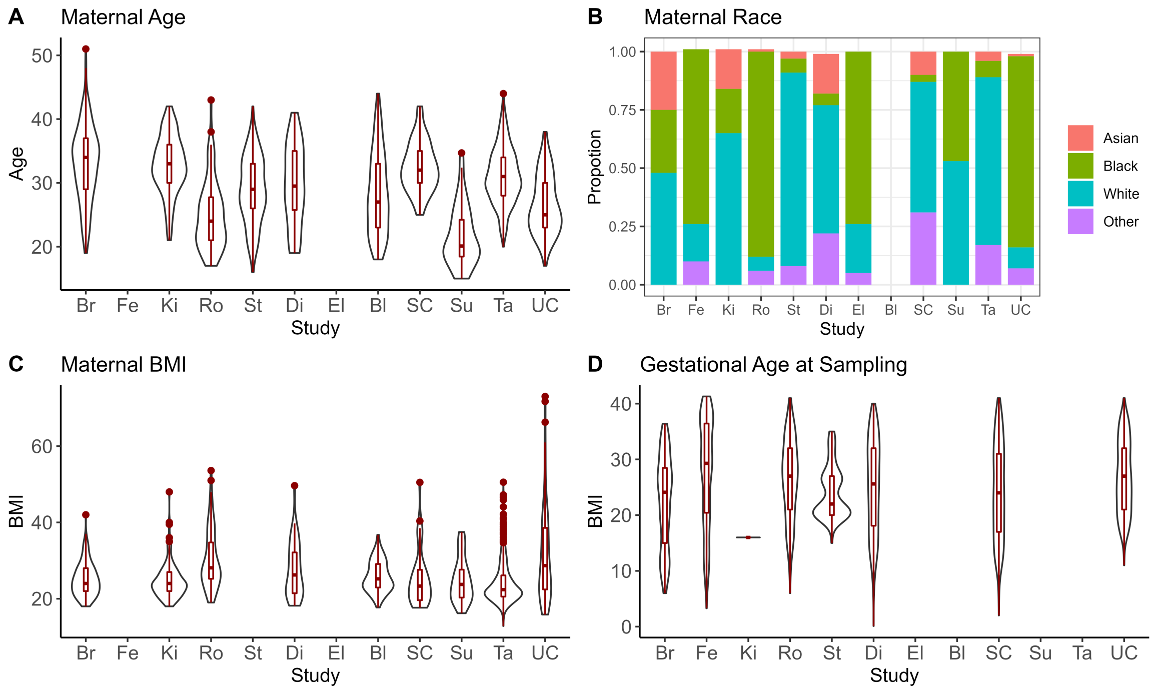


**Figure S2.** The summary statistics of the population characteristics and gestational age at sampling.

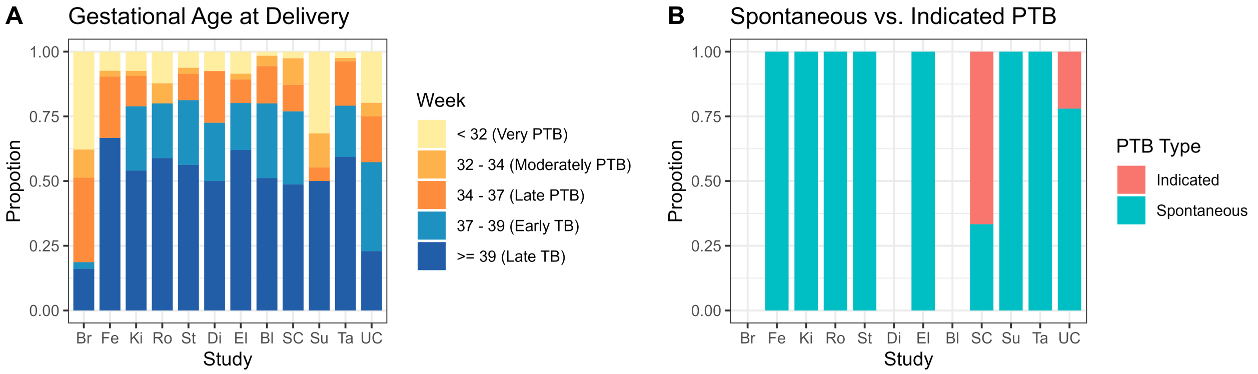


**Figure S3.** A: Distribution of gestational age delivery; B: Preterm birth type inlucded in each dataset.

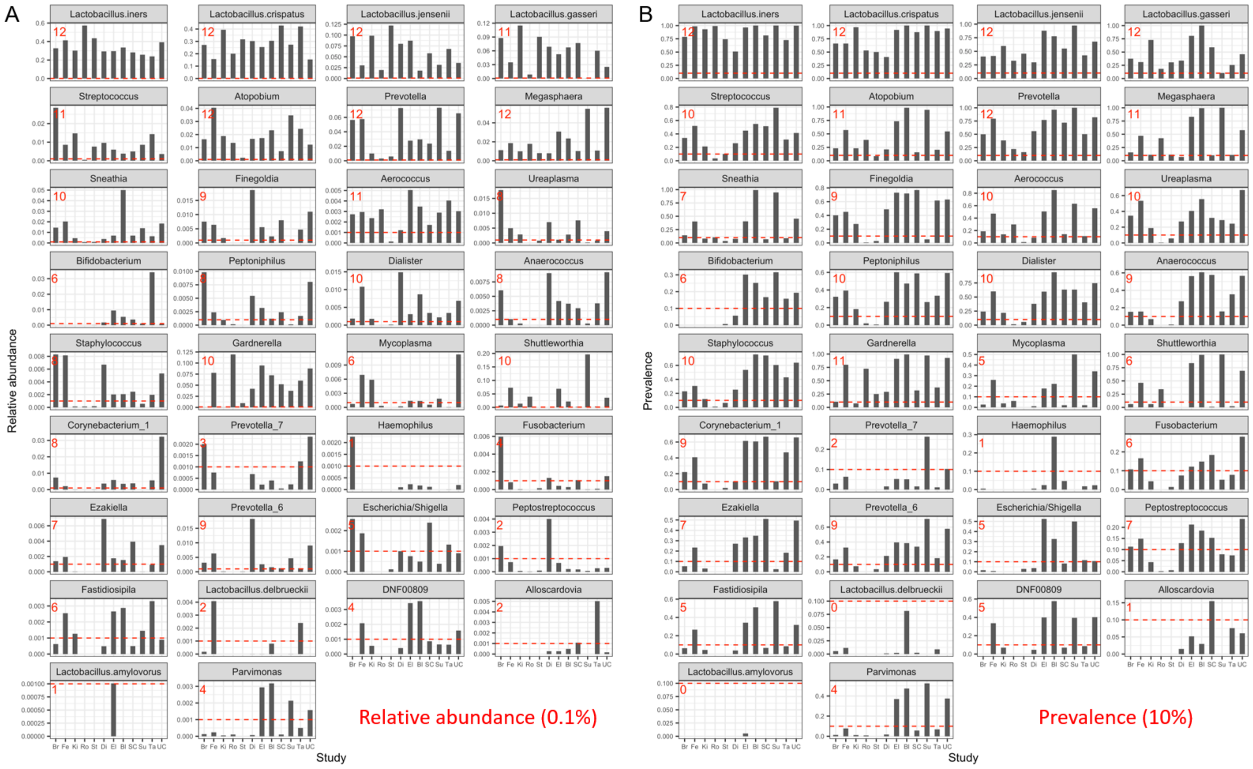


**Figure S4.** The relative abundance (A) and prevalence (B) of 34 genera/species either in common or top feature tables for each dataset. The red number in the up-left shows the number of datasets that have relative abundance greater than 0.1% or prevalence greater than 10%. After filtering the genera using the following criteria (1) at least 5 datasets have the average relative abundance > 0.1%; (2) at least 5 datasets have the average prevalence > 10%, we excluded Prevotella_7, Haemophilus, Lactobacillus.delbrueckii, Alloscardovia, Lactobacillus.amylovorus, Fusobacterium, Peptostreptococcus, DNF00809, Parvimonas and kept another 25 genera/species for further analysis.

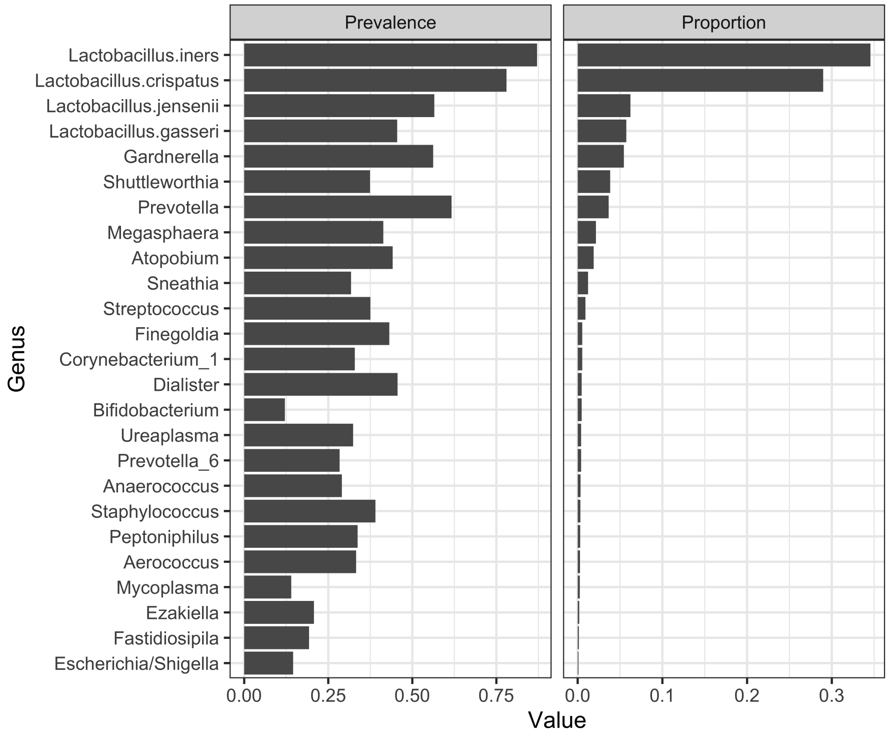


**Figure S5.** The average prevalence and relative abundance of 25 core taxa across datasets

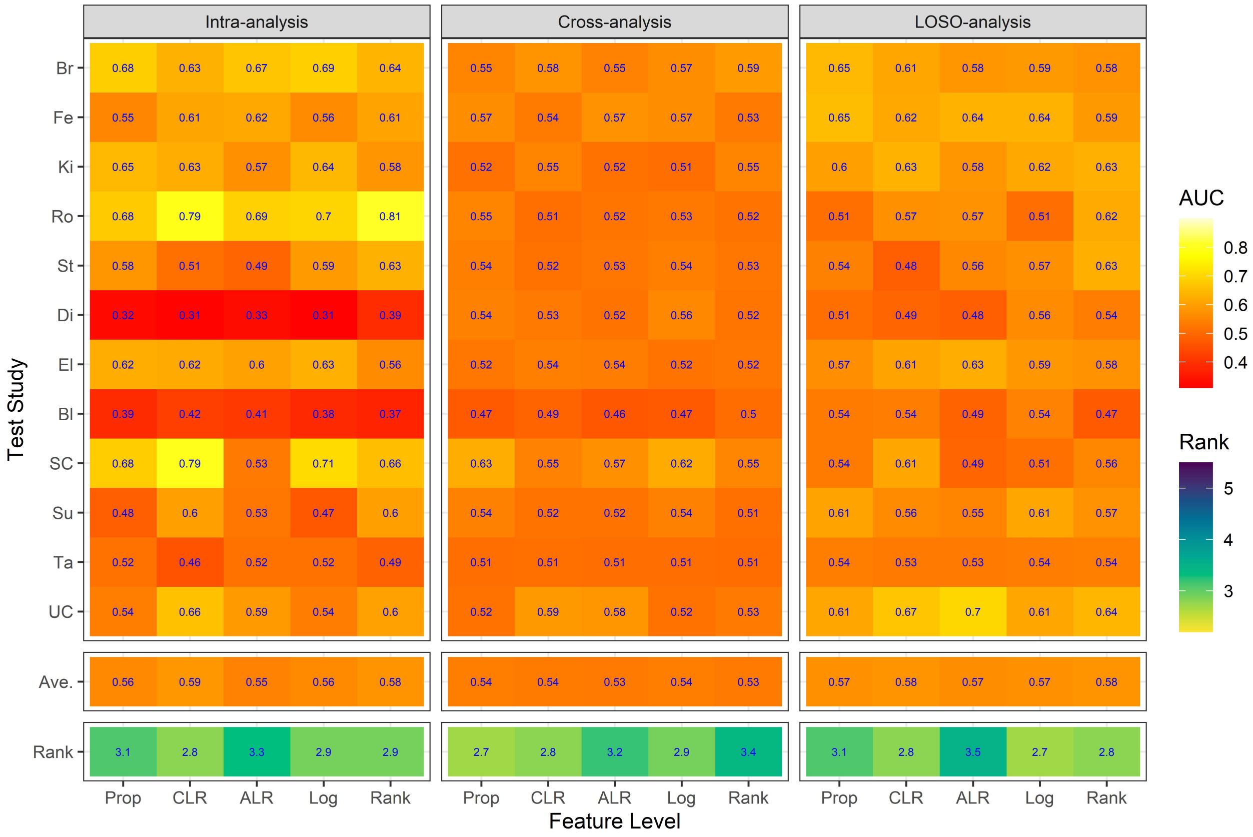


**Figure S6.** Assessment prediction performance of data transformation methods using the genus features. For intra-analysis, the area under the receiver operating characteristics curve (AUC) is obtained based on cross-validation within a dataset. For cross-analysis, the AUC value for a given dataset is the average of the AUCs using other dataset in V1-V2 or V4 group as the training set, and the dataset as the testing set. For LOSO-analysis, the AUC value for a given dataset is obtained using the combined dataset in V1-V2 or V4 group except the dataset as training set and the study as the testing set. Rank is calculated as the average rank of feature levels in terms of the AUC value for each study. Larger AUC has a smaller rank.

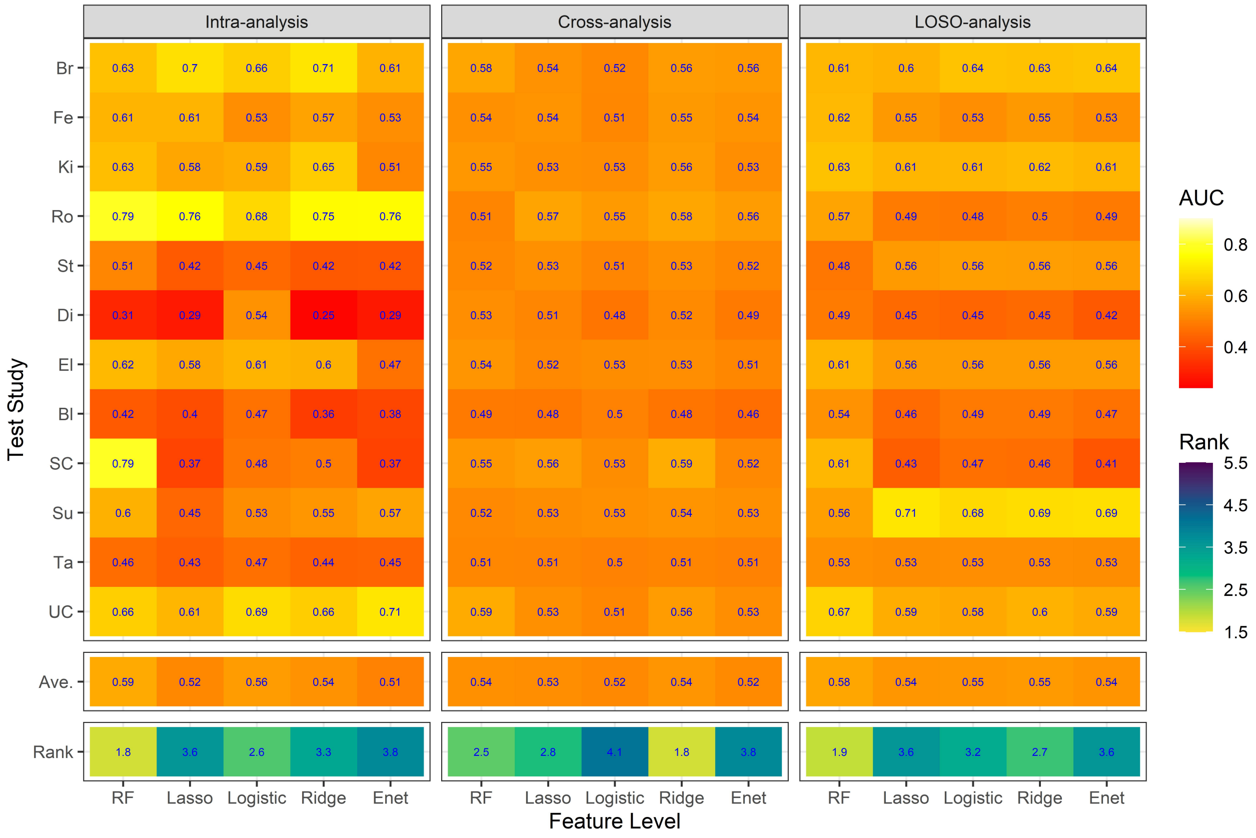


**Figure S7.** Assessment prediction performance of classifiers using the genus features with CLR transformation.

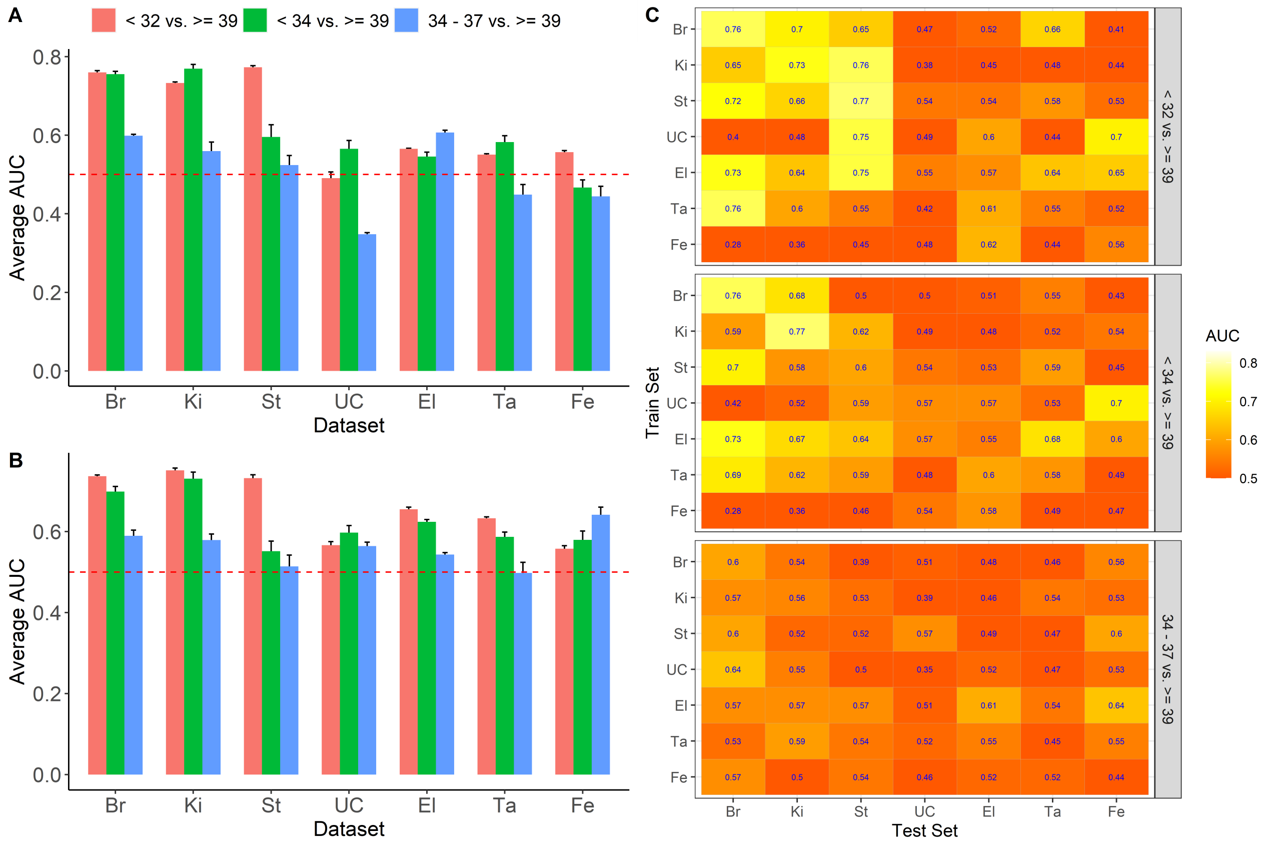


**Figure S8.** Assessment of prediction performance for different preterm birth groups using intra-dataset analysis (A), combined-dataset analysis (B) and cross-dataset analysis (C) using proportional abundance data.

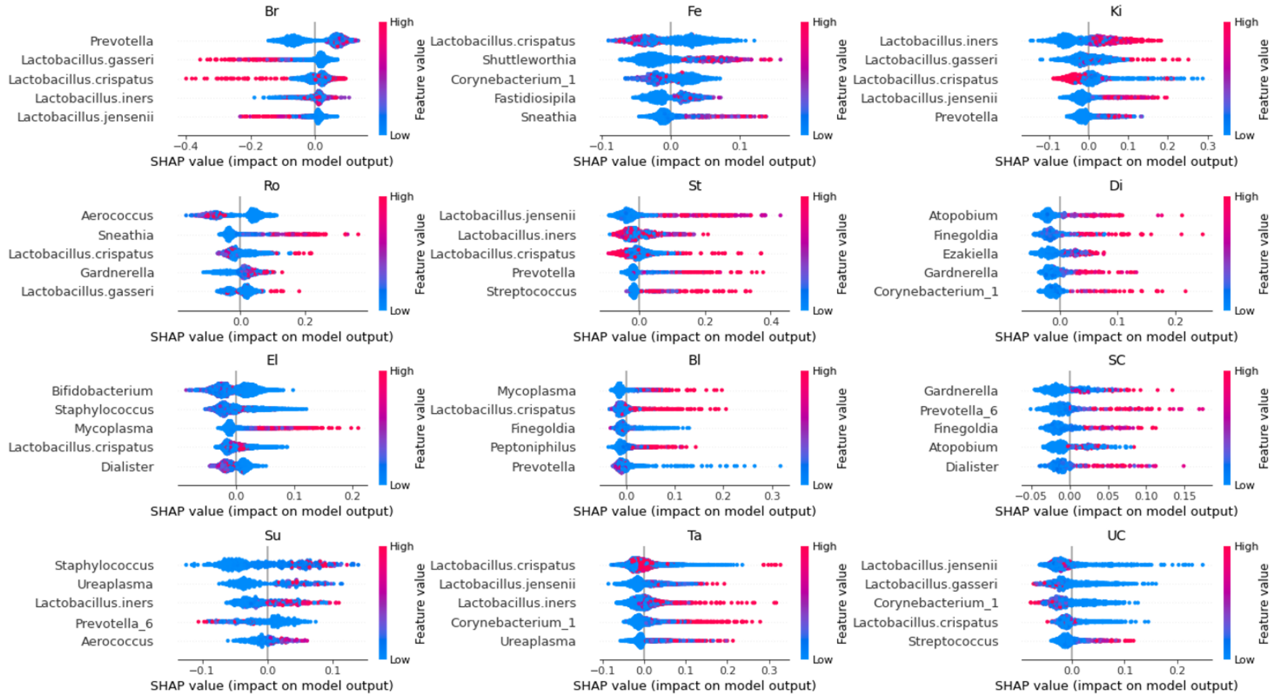


**Figure S9.** SHAP summary plots for each study for random forests trained on genus-level proportional data. The five most important features for each study are shown. The data points are colored by the relative abundance of the taxa. Positive SHAP values indicate a higher likelihood of preterm birth and negative SHAP values indicate a lower likelihood of preterm birth in comparison to the average prediction.

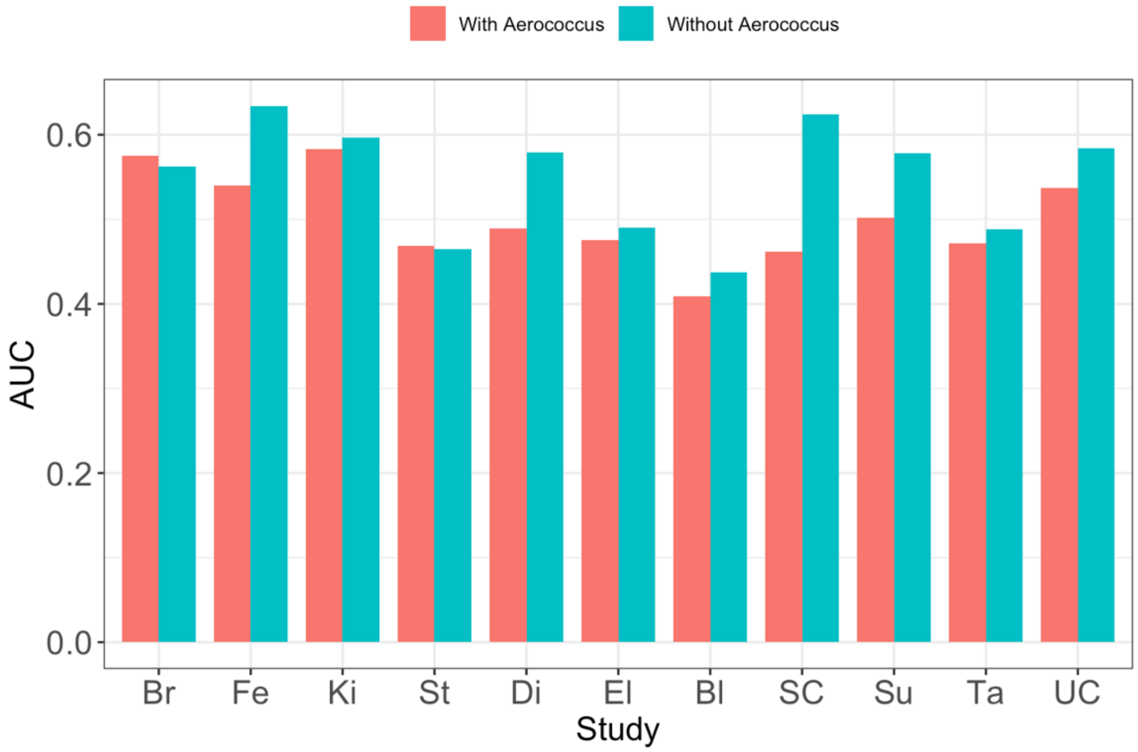


**Figure S10.** Average AUC with and without Aerococcus included as a feature for ten random forest models trained on genus-level CLR data from the Ro study and tested on all other studies.

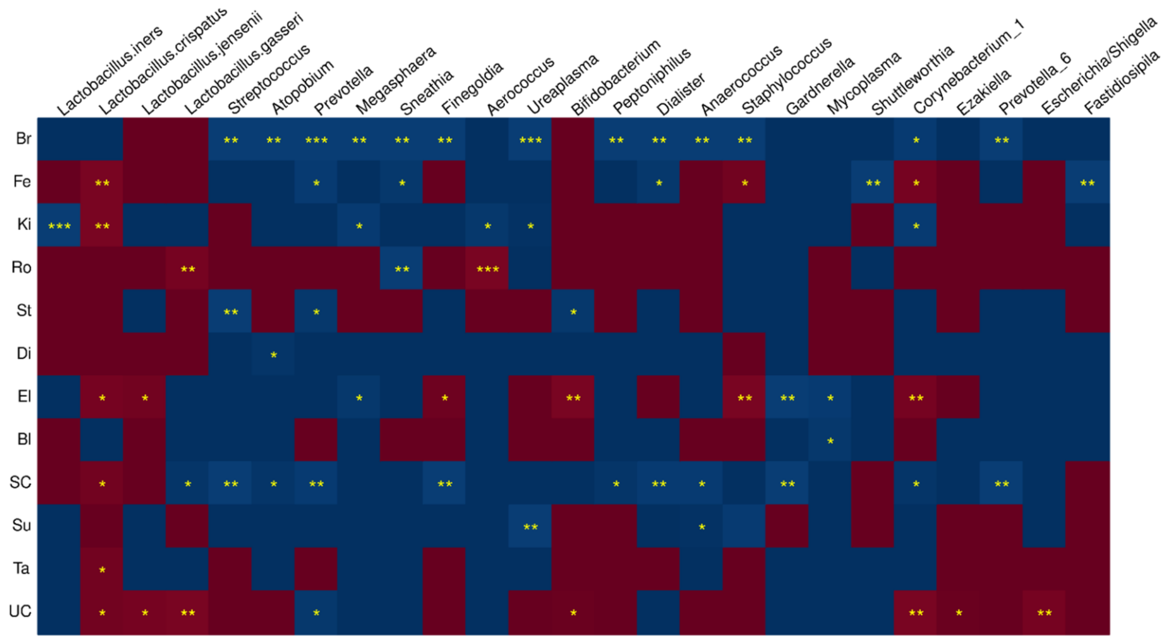


**Figure S11.** Dataset-specific differential abundance analysis for each genus/species using one-sided Wilcoxon rank-sum test. A cell is in red color if the average relative abundance of a genus/species in term birth is larger than the average relative abundance in preterm birth, otherwise in blue color. The unadjusted p-value significant code: '***': 0.001, '**': 0.01, '*': 0.05.

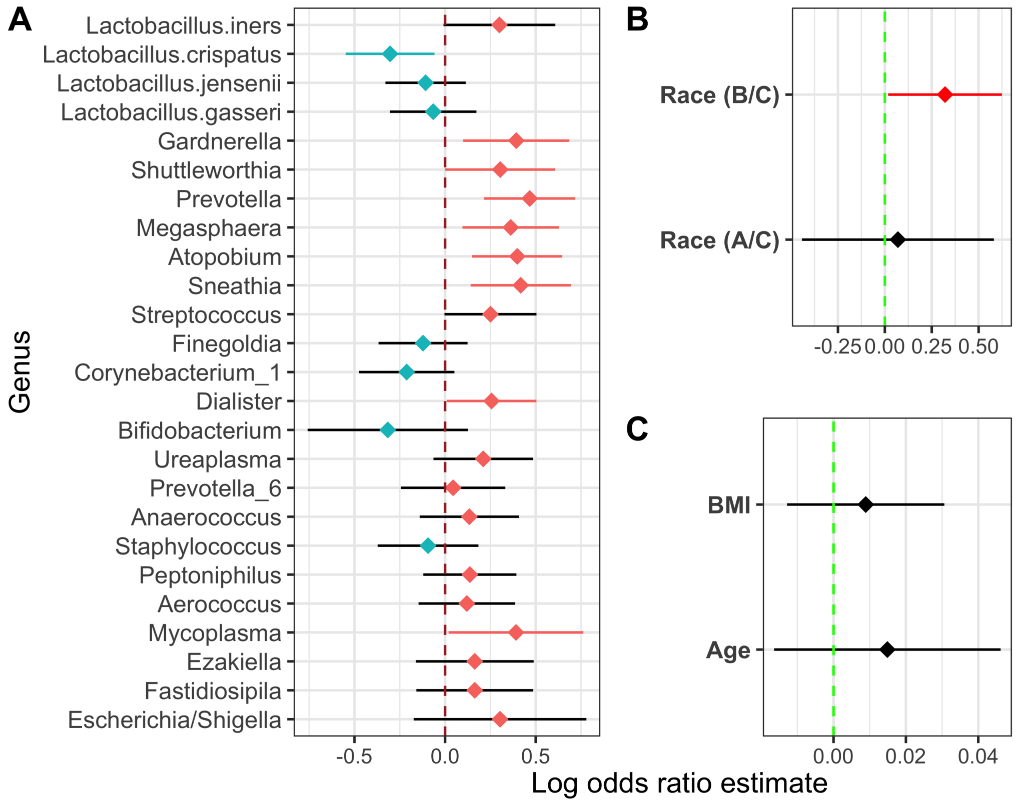


**Figure S12.** Cross-dataset differential abundance analysis using a generalized linear mixed model with covariates. The genus and race effects were estimated using 11 datasets with a model including race covariate. The BMI and age effects were estimated using 8 datasets with a model including race, BMI and age. Point estimates less than 0 are shown as blue points and greater than 0 are shown as red points. Confident intervals less than 0 are shown as blue bars and greater than 0 are shown as red bars.

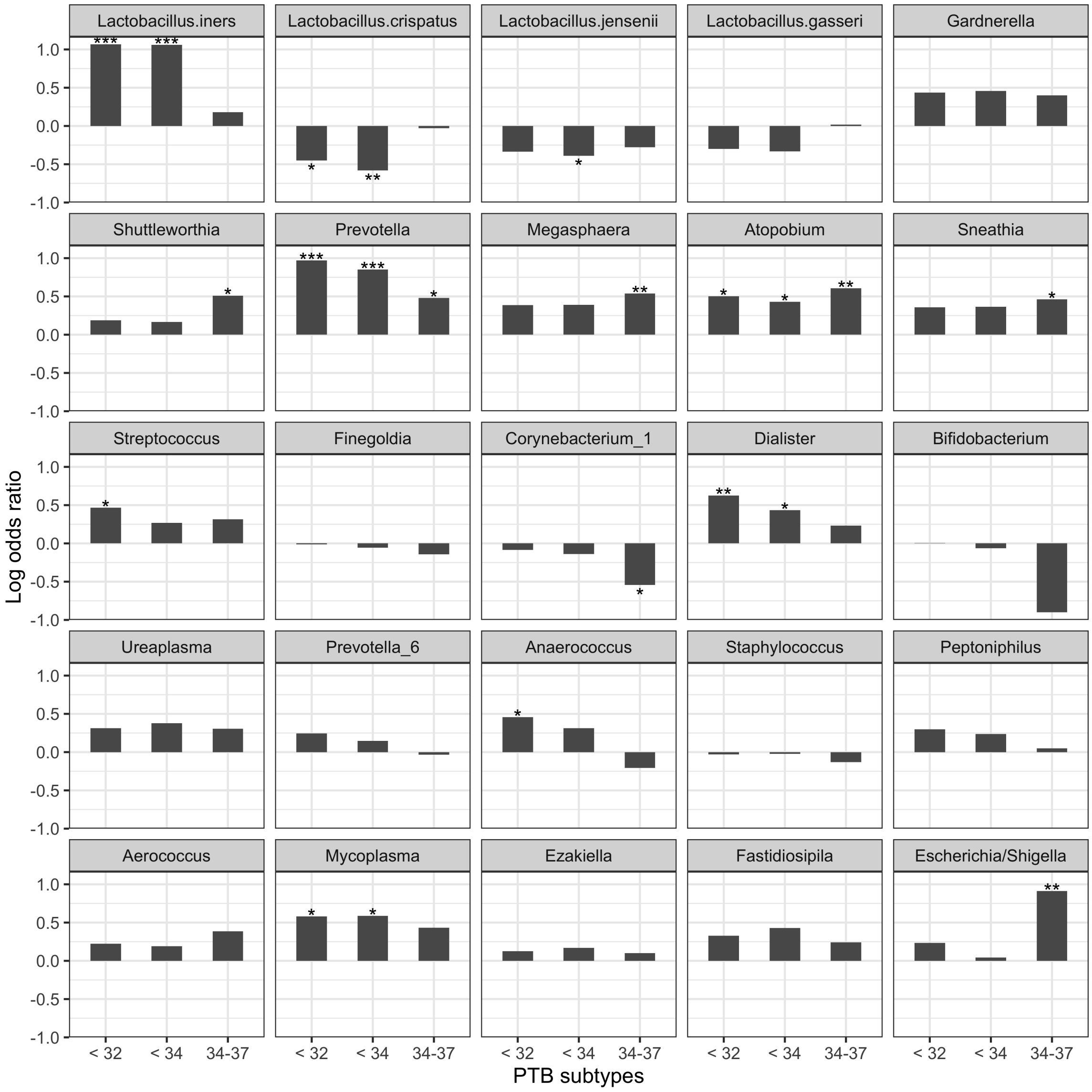


**Figure S13.** Average estimates of log odd ratio for each PTB subgroup versus late term birth using the generalized linear mixed model. The asterisk shows the significant level of the combined p-values: *: p-value < 0.05; **: p-value < 0.01; ***: p-value < 0.001.

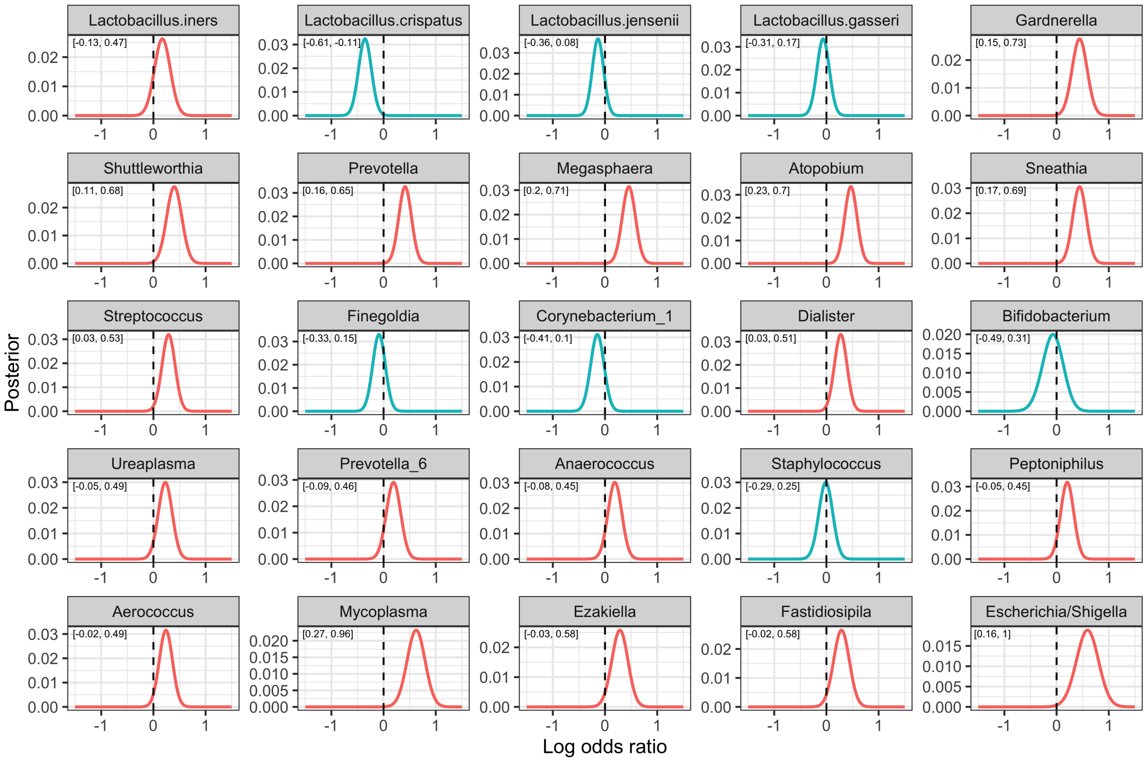


**Figure S14.** Posterior distribution of log odds ratio using a Bayesian approach stepwise method. Maximum a posteriori (MAP) estimates less than 0 are shown as blue lines and larger than 0 are shown as red lines. The 95% credible intervals are shown at the left top corner.

#### Supplementary Tables

**Table S1.** Number of subjects that have samples from two or more trimesters.

| Trimester | Br | Fe | Ki | Ro | St | Di | El | Bl | SC | Su | Ta | UC |
|---|---|---|---|---|---|---|---|---|---|---|---|---|
| first & second | 40 | 46 | 0 | 12 | 0 | 23 | 0 | 0 | 36 | 0 | 0 | 3 |
| first & third | 20 | 50 | 0 | 7 | 0 | 21 | 0 | 0 | 36 | 0 | 0 | 3 |
| second & third | 27 | 93 | 0 | 49 | 13 | 33 | 0 | 0 | 39 | 0 | 0 | 84 |
| all three | 17 | 45 | 0 | 7 | 0 | 21 | 0 | 0 | 36 | 0 | 0 | 3 |

**Table S2.** Data available for the datasets included in this meta-analysis

| Dataset | Sequence data | Metadata |
|---|---|---|
| Brown2018 (Br)* | ENA (PRJEB30642 and PRJEB21325) | From authors |
| Fettweis2019 (Fe) | SRA (PRJNA326441) | dbGap (phs001523) |
| Kindinger2017 (Ki) | ENA (PRJEB11895 and PRJEB12577) | From authors |
| Romero2014 (Ro) | SRA (PRJNA242473) | SRA (PRJNA242473) |
| Stafford2017 (St) | SRA (SRP065627) | Suppl. material |
| Digiulio2015 (Di) | SRA (SRP288562)) | From authors |
| Elovitz2019 (El) | dbGap (phs001739) | dbGap (phs001739 |
| Blostein2020 (Bl) | From authors | From authors |
| ST_Callahan2017 (SC)** | Suppl. material | Suppl. material |
| Subramaniam2018 (Su) | SRA (PRJNA600021) | SRA (PRJNA600021) |
| Tabatabaei2019 (Ta) | From authors | From authors |
| UAB_Callahan2017 (UC)** | Suppl. material | Suppl. material |

* Dataset Brown2018 was combined from Brown et al. (2018) and Brown et al. (2019).

** For Callahan2017, DADA2 is used to processed sequence data.

**Table S3.** Additional16S rRNA sequencing data information for the datasets included in this meta-analysis

| Dataset | Sequencing Platform (Region) | Primer | If paired | Which used? |
|---|---|---|---|---|
| Brown2018 (Br)* | Illumina Miseq (V1-V2) | 28F/388R | Yes | Forward |
| Fettweis2019 (Fe) | Illumina Miseq (V1-V3) | - | Yes | Forward |
| Kindinger2017 (Ki) | Illumina Miseq (V1-V3) | 28F/519F | Yes | Forward |
| Romero2014 (Ro) | 454 amplicon Seq (V1-V3) | 27F/534R | No | Forward |
| Stafford2017 (St) | 454 amplicon Seq (V1-V3) | 27F/519R | No | Forward |
| Digiulio2015 (Di) | 454 amplicon Seq (V3-V5) | 338F/906R | No | Forward |
| Elovitz2019 (El) | Illumina Hiseq (V3-V4) | 319F/806R | Yes | Forward and reverse |
| Blostein2020 (Bl) | Illumina Treseq (V4) | 515F/806R | Yes | Forward and reverse |
| ST_Callahan2017 (SC) | Illumina Hiseq (V4) | 515F/806R | Yes | Forward and reverse |
| Subramaniam2018 (Su) | Illumina Miseq (V4) | 515F/806R | Yes | Forward |
| Tabatabaei2019 (Ta)* | Illumina (V4) | 515F/806R | - | - |
| UAB_Callahan2017 (UC) | Illumina Hiseq (V4) | 515F/806R | Yes | Forward and reverse |

* For Tabatabaei2019, only the processed FASTA file is available to us. We re-created the FASTQ file using an arbitrary quality score.

**Table S4.** Number of features at ASV and other taxonomic levels.

| Feature Level | V1-V2 group | | V4 group | |
|---|---|---|---|---|
| | Common | Top | Common | Top |
| Phylum | 6 | 8 | 9 | 10 |
| Class | 9 | 12 | 15 | 14 |
| Order | 10 | 20 | 24 | 23 |
| Family | 13 | 26 | 42 | 35 |
| Genus | 22 | 45 | 84 | 63 |
| ASV | 42 | 172 | 157 | 159 |

**Table S5.** Average AUC of using different feature levels using common feature table for early PTB (<32 weeks) and early or moderate PTB (<34 weeks) subgroups.

| PTB groups | Analysis | V1-V2 group (4 datasets) | | | | | | V4 group (3 datasets) | | | | | |
|---|---|---|---|---|---|---|---|---|---|---|---|---|---|
| | | ASV | Genus | Family | Order | Class | Phylum | ASV | Genus | Family | Order | Class | Phylum |
| <32 weeks | Intra | 0.71 | 0.70 | 0.67 | 0.68 | 0.69 | 0.66 | 0.63 | 0.60 | 0.65 | 0.63 | 0.58 | 0.60 |
| | Cross | 0.59 | 0.60 | 0.52 | 0.51 | 0.52 | 0.51 | 0.56 | 0.60 | 0.59 | 0.60 | 0.56 | 0.59 |
| | LODO | 0.66 | 0.68 | 0.58 | 0.58 | 0.61 | 0.62 | 0.58 | 0.61 | 0.62 | 0.62 | 0.57 | 0.63 |
| <34 weeks | Intra | 0.65 | 0.63 | 0.62 | 0.62 | 0.62 | 0.61 | 0.63 | 0.62 | 0.65 | 0.63 | 0.61 | 0.61 |
| | Cross | 0.54 | 0.55 | 0.52 | 0.51 | 0.51 | 0.50 | 0.56 | 0.59 | 0.57 | 0.57 | 0.55 | 0.58 |
| | LODO | 0.60 | 0.61 | 0.55 | 0.56 | 0.58 | 0.62 | 0.57 | 0.58 | 0.57 | 0.58 | 0.57 | 0.58 |

**Table S6.** Estimate of the odds ratio between genus is present relative to absent.

| Genus/Species | Estimate | 95% CI | p-value | adjusted p-value |
|---|---|---|---|---|
| Lactobacillus.iners | 1.37 | [1.01, 1.87] | 0.0425 | 0.101 |
| Lactobacillus.crispatus | 0.72 | [0.56, 0.91] | 0.0070 | 0.029 |
| Lactobacillus.jensenii | 0.86 | [0.70, 1.07] | 0.1723 | 0.273 |
| Lactobacillus.gasseri | 0.91 | [0.72, 1.15] | 0.4080 | 0.425 |
| Gardnerella | 1.56 | [1.18, 2.08] | 0.0021 | 0.013 |
| Shuttleworthia | 1.40 | [1.06, 1.87] | 0.0199 | 0.062 |
| Prevotella | 1.57 | [1.23, 1.99] | 0.0002 | 0.003 |
| Megasphaera | 1.54 | [1.20, 1.98] | 0.0007 | 0.006 |
| Atopobium | 1.58 | [1.25, 1.99] | 0.0001 | 0.003 |
| Sneathia | 1.48 | [1.15, 1.92] | 0.0027 | 0.014 |
| Streptococcus | 1.29 | [1.01, 1.65] | 0.0443 | 0.101 |
| Finegoldia | 0.89 | [0.70, 1.12] | 0.3208 | 0.382 |
| Corynebacterium_1 | 0.81 | [0.62, 1.04] | 0.0947 | 0.197 |
| Dialister | 1.30 | [1.03, 1.65] | 0.0302 | 0.084 |
| Bifidobacterium | 0.70 | [0.45, 1.08] | 0.1041 | 0.200 |
| Ureaplasma | 1.18 | [0.91, 1.54] | 0.2128 | 0.305 |
| Prevotella_6 | 1.13 | [0.86, 1.48] | 0.3863 | 0.420 |
| Anaerococcus | 1.13 | [0.87, 1.46] | 0.3764 | 0.420 |
| Staphylococcus | 0.92 | [0.71, 1.20] | 0.5406 | 0.541 |
| Peptoniphilus | 1.17 | [0.91, 1.50] | 0.2196 | 0.305 |
| Aerococcus | 1.21 | [0.94, 1.55] | 0.1402 | 0.250 |
| Mycoplasma | 1.60 | [1.13, 2.28] | 0.0081 | 0.029 |
| Ezakiella | 1.18 | [0.86, 1.61] | 0.3094 | 0.382 |
| Fastidiosipila | 1.18 | [0.87, 1.60] | 0.2796 | 0.368 |
| Escherichia/Shigella | 1.36 | [0.87, 2.14] | 0.1744 | 0.273 |

### Bibliography

Brown, R. G., Marchesi, J. R., Lee, Y. S., Smith, A., Lehne, B., Kindinger, L. M., Terzidou, V., Holmes, E., Nicholson, J. K., Bennett, P. R., et al. Vaginal dysbiosis increases risk of preterm fetal membrane rupture, neonatal sepsis and is exacerbated by erythromycin. *BMC medicine*, 16(1):1–15, 2018.

Brown, R. G., Al-Memar, M., Marchesi, J. R., Lee, Y. S., Smith, A., Chan, D., Lewis, H., Kindinger, L., Terzidou, V., Bourne, T., et al. Establishment of vaginal microbiota composition in early pregnancy and its association with subsequent preterm prelabor rupture of the fetal membranes. *Translational Research*, 207:30–43, 2019.

### Supplementary Methods

#### Classifiers

We implemented the following classifiers in the machine learning framework and compared their performance: (1) random forest; (2) logistic regression; (3) L1-regularized (LASSO) logistic regression; (4) L2-regularized (Ridge) logistic regression; (5) Elastic net regularized logistic regression. Random forest was implemented using the randomForest R package with 1000 trees. The hyperparameter 'mtry' (number of variables randomly sampled as candidates at each split) was tuned using the caret R package with repeated cross-validation. Logistic regression was implemented using the glm function in R. The regularized logistic regression models were implemented using the glmnet R package. The hyperparameter 'lambda' in the LASSO and Ridge models was tuned using the cv.glment function. The hyperparameters 'alpha' and 'lambda' were tuned using the caret R package with repeated cross-validation.

#### Bayesian Approach

Let $G = 1$ is a genus is present (relative abundance $> 0.001$) and $0$ if absent, $B = 1$ if a women has preterm birth (PTB) and $0$ if term birth. We can have the conditional probability of PTB giving a genus is absent for dataset $i$ to be

$$p_i(B = 1|G = 0) = p_i^0 = \frac{u_i}{1+u_i}$$

where $u_i$ is the odds of PTB giving a genus is absent for dataset $i$.
Define $r$ as the odds ratio between a genus is present and absent and it is the same for different datasets. Thus, the conditional probability of PTB giving a genus is present for dataset $i$ can be written as

$$p_i(B = 1|G = 1) = p_i^1 = \frac{u_i r}{1+u_i r}$$

We assume both $u_i$ and $r$ have prior distributions, $p(u_i)$ and $p(r)$, respectively. We are interested in calculating the posterior distribution of $r$. Given dataset $i$, let $N_i$ is the total number of subjects in study $i$, $n_i$ is the number of subjects that with certain genus present. $M_i$ is the number of PTB subjects in study $i$, $m_i$ is the number of PTB subjects that with certain genus present. Thus, we can have the likelihood function

$$\begin{aligned} & p(N_i, n_i, M_i, m_i | u_i, r) \\ & = (p_i^0)^{M_i - m_i}(1-p_i^0)^{(N_i-n_i)-(M_i-m_i)}(p_i^1)^{m_i}(1-p_i^1)^{n_i-m_i} \\ & = \frac{u_i^{M_i-m_i}}{(1+u_i)^{N_i-n_i}} * \frac{(u_i r)^{m_i}}{(1+u_i r)^{n_i}} \\ & = \frac{u_i^{M_i} r^{m_i}}{(1+u_i)^{N_i-n_i}(1+u_i r)^{n_i}} \end{aligned} \qquad \textbf{(S1)}$$

Thus, the posterior distribution of $u_i$ and $r$ can be written as

$$\begin{aligned} p(u_i, r | N_i, n_i, M_i, m_i) &= \frac{p(N_i, n_i, M_i, m_i | u_i, r) p(u_i, r)}{p(N_i, n_i, M_i, m_i)} \\ &= \frac{p(N_i, n_i, M_i, m_i | u_i, r) p(u_i) p(r)}{\int_{u_i} \int_r p(N_i, n_i, M_i, m_i | u_i, r) p(u_i) p(r)} \end{aligned} \qquad \textbf{(S2)}$$

Furthermore, we can integrate out $u_i$ and obtain the posterior distribution for odds ratio $r$,

$$\begin{aligned} p(r | N_i, n_i, M_i, m_i) &= \int_{u_i} p(u_i, r | N_i, n_i, M_i, m_i) \\ &= \frac{p(r) \int_{u_i} p(N_i, n_i, M_i, m_i | u_i, r) p(u_i)}{\int_{u_i} \int_r p(N_i, n_i, M_i, m_i | u_i, r) p(u_i) p(r)} \end{aligned} \qquad \textbf{(S3)}$$

We assume the $\log(u_i)$ follows a uniform prior distribution for each dataset. For $r$, we let the first dataset has the uniform prior distribution, then calculate the posterior distribution of $r$. Next we let the posterior distribution of the odds ratio from the first dataset be the prior distribution for the second dataset, and update the posterior distribution of $r$. Repeated the process until the last dataset and obtain the final posterior distribution of $r$.
